# GWAS of ∼30,000 samples with bone mineral density at multiple skeletal sites and its clinical relevance on fracture prediction, genetic correlations and prioritization of drug targets

**DOI:** 10.1101/2024.01.18.24301465

**Authors:** Yu Qian, Jiangwei Xia, Pingyu Wang, Chao Xie, Hong-Li Lin, Gloria Hoi-Yee Li, Cheng-Da Yuan, Mo-Chang Qiu, Yi-Hu Fang, Chun-Fu Yu, Xiang-Chun Cai, Saber Khederzadeh, Pian-Pian Zhao, Meng-Yuan Yang, Jia-Dong Zhong, Xin Li, Peng-Lin Guan, Jia-Xuan Gu, Si-Rui Gai, Xiang-Jiao Yi, Jian-Guo Tao, Xiang Chen, Mao-Mao Miao, Guo-Bo Chen, Lin Xu, Shu-Yang Xie, Geng Tian, Hua Yue, Guangfei Li, Wenjin Xiao, David Karasik, Youjia Xu, Liu Yang, Ching-Lung Cheung, Fei Huang, Zhenlin Zhang, Hou-Feng Zheng

## Abstract

We conducted genome-wide association studies (GWAS) of dual-energy X-ray absorptiometry (DXA)-derived bone mineral density (BMD) traits at 11 skeletal sites, within over 30,000 European individuals from the UK Biobank. A total of 92 unique and independent loci were identified for 11 DXA-derived BMD traits and fracture, including five novel loci (harboring genes such as *ABCA1*, *CHSY1*, *CYP24A1*, *SWAP70*, and *PAX1*) for six BMD traits. These loci exhibited evidence of association in both males and females, which could serve as independent replication. We demonstrated that polygenic risk scores (PRSs) were independently associated with fracture risk. Although incorporating multiple PRSs (metaPRS) with the clinical risk factors (i.e., the FRAX model) exhibited the highest predictive performance, the improvement was marginal in fracture prediction. The metaPRS were capable of stratifying individuals into different trajectories of fracture risk, but clinical risk factors played a more significant role in the stratification. Additionally, we uncovered genetic correlation and shared polygenicity between head BMD and intracranial aneurysm. Finally, by integrating gene expression and GWAS datasets, we prioritized genes (e.g. *ESR1* and *SREBF1*) encoding druggable human proteins along with their respective inhibitors/antagonists. In conclusion, this comprehensive investigation revealed a new genetic basis for BMD and its clinical relevance on fracture prediction. More importantly, it was suggested that head BMD was genetically correlated with intracranial aneurysm. The prioritization of genetically supported targets implied the potential repurposing drugs (e.g. the n-3 PUFA supplement targeting SREBF1) for the prevention of osteoporosis.

## Introduction

Osteoporosis, a systemic skeletal disease characterized by decreased bone mass and micro-structural damage ^1,2^, has a global prevalence of 18.3% [95% confidence interval (95% CI): 16.2%-20.7%] ^3^. Bone mass could be assessed by 2-dimensional projectional scans with dual-energy X-ray absorptiometry (DXA), or other medical imaging tools, such as quantitative computed tomography (QCT) and quantitative ultrasound (QUS) ^4^. Genome-wide association studies (GWASs) and meta-analyses were carried out to explore the genetic factors for bone mineral density (BMD), osteoporosis, and fracture ^1,2^. Early GWAS design only involved thousands of samples and only several loci were identified ^5,6^. The meta-analysis could enlarge the sample size and statistical power, and lead to the identification of more loci ^7,8^. However, the genetic summary data, instead of individual-level genotype data, from each cohort were meta-analyzed in the aforementioned studies. Recently, large-scale biobanks such as the UK biobank could enable access to the individual-level genotype data in hundreds of thousands of samples, and hundreds of genetic loci were identified for QUS-derived BMD in these efforts ^9,10^.

Although GWASs have been successfully conducted in the past decade, the ultimate goal of genetic study is to translate the discoveries into clinical practice. Previously, we have tried to summarize the clinical use of GWAS findings in the bone field, such as disease prediction ^1^. Lu et al developed the genetically predicted speed of sound (SOS, a parameter measured by QUS) for individuals in UK Biobank by common genetic variants through polygenic risk score (PRS) ^11^. They demonstrated that this score provided modestly better fracture risk prediction than some of the clinical risk factors such as smoking and use of corticosteroids ^11^. In addition, they suggested that adding rare variants did not demonstrate substantially improved predictive performance in a recent study ^12^. The above studies took the SOS measurement in the training and testing dataset, however, the SOS measurement was not correlated very well with BMD ^13^. Another clinical relevance of GWAS findings is to infer the correlation between diseases ^1^. Earlier efforts have uncovered numerous SNPs exhibiting pleiotropic associations with BMD and other traits/diseases, such as birth weight ^14^, type 2 diabetes ^15^, and major depressive disorder ^16^. Finally, incorporating genetic data in drug development is warranted to improve this process, because drugs with genetic support are more likely to succeed in clinical trials ^17,18^.

Therefore, with the availability of DXA-derived BMD phenotypes and individual-level genotype data in UK Biobank, it is an opportunity to conduct a genome-wide association study at large scale individual-level genotype data and to investigate the genetic basis of BMD at 11 sites (arm, femur total, femur neck, head, leg, pelvis, lumbar spine, rib, and spine) and fracture (**Supplementary Figure 1**). We then build a ‘multi-BMD PRS’ predictive model to improve genetic risk stratification for fracture. In addition, we estimated the shared genetic architecture of BMD with other common chronic diseases, including neurodegenerative, cardiovascular, and autoimmune diseases. Finally, we tried to explore the potential effective and safe therapeutic targets for osteoporosis.

## Results

### Genetic architecture of BMD at multiple skeletal sites

The overview of the study design was presented in Supplementary Figure 1. Specifically, we conducted the GWAS analyses for BMD at 11 anatomic sites (i.e., arm, total femur, femoral neck, head, legs, lumbar spine, pelvis, ribs, spine, trunk, and total body) and any-type fracture (**Figure 1A**) in male and female separately. For each BMD trait, we then conducted meta-analyses to combine the results from both genders (for BMD traits: N≈30,000; for any-type fracture: N=35,192 for cases; N=317,599 for controls). The reported loci should exhibit evidence of association in both males and females, which could serve as independent replication (**Figure 2 A-E Supplementary Table 1,** and **Supplementary Table 2**). All intercept values from the LD score method were close to one, revealing no obvious population stratification for all GWASs (**Supplementary Table 3**). We observed that approximately 25.7%∼41.8% of the variance in BMD and 4.8% of the variance in fracture risk could be explained by common variants across the genome (**Table 1**). We then conducted conditional analyses within phenotype and identified 240 unique conditional independent BMD signals (**Table 1, Figure 1B, Supplementary Table 1**, and **Supplementary Figure 2-13**). After merging the physically overlapped signals across BMD phenotypes (i.e., the distance between two conditional independent SNVs < 500kb) into one locus, a total of 91 unique and independent BMD loci were defined (**Table 1, Figure 1B** and **Supplementary Table 1**). We identified 8 loci for fracture, 7 of which overlapped with the above BMD signals, and one of which (independent SNP: rs13281992) was previously reported to be genome-wide significant associated with heel BMD ^10^ (**Table 1, Figure 1B** and **Supplementary Table 1**).

**Figure 1.**
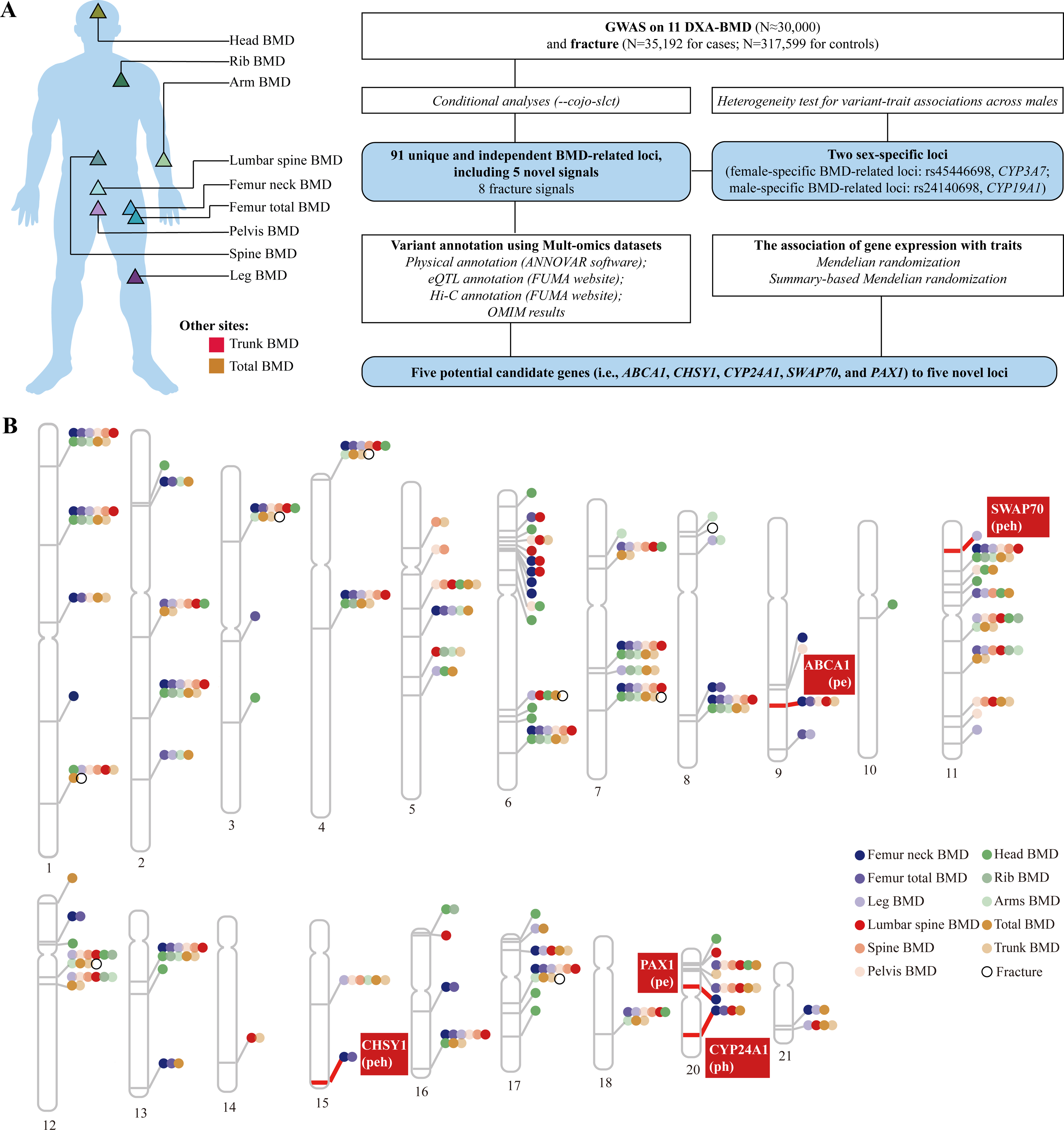
Genetic architecture of DXA-BMD at multiple skeletal sites. (A) the skeletal sites of 11 DXA-derived BMD traits and the GWAS study design; (B) the associated genetic loci for DXA-derived BMD and fracture; Abbreviations: DXA, dual-energy X-ray absorptiometry; BMD, bone mineral density; GWAS, genome-wide association study.

**Figure 2.**
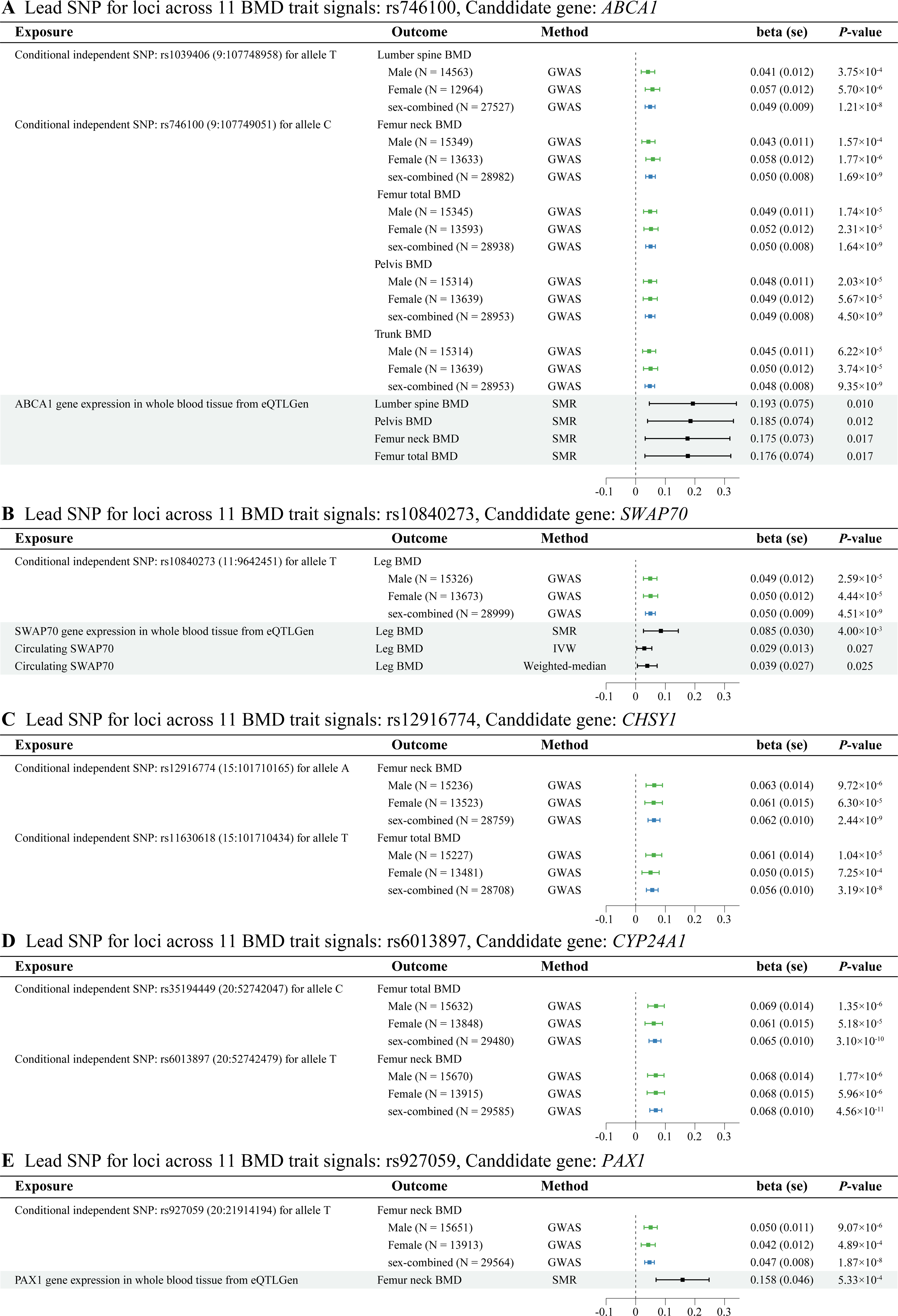
Forest plot of the genetic association estimates of lead SNPs (A: rs746100; B: rs10840273; C: rs12916774; D: rs6013897; E: rs927059) of novel loci with corresponding BMD traits in female GWASs and male GWASs as well as GWAS meta-analyses of both sexes, along with the result from Mendelian randomization analyses. Abbreviations: BMD, bone mineral density; eQTL, expression quantitative trait locus; IVW, inverse variance weighted; GWAS, genome-wide association study; pQTL, genotype–protein association; SMR, summary-based Mendelian randomization.

**Table 1.** the detailed information on genome-wide association studies of 11 bone mineral density sites.

| Phenotype | Sample size | Conditional independent signals |  |  | Heritability |
| --- | --- | --- | --- | --- | --- |
|  |  | Known | Novel | Total |  |
| **Single BMD phenotype** |  |  |  |  |  |
| Head BMD | 31986 | 69 | 0 | 69 | 0.418 (0.045) |
| Arm BMD | 31873 | 34 | 0 | 34 | 0.321 (0.039) |
| Femoral neck BMD | 32017 | 36 | 4 | 40 | 0.257 (0.029) |
| Femur total BMD | 31873 | 41 | 3 | 44 | 0.319 (0.030) |
| Leg BMD | 31873 | 43 | 1 | 44 | 0.320 (0.034) |
| Lumbar spine BMD | 30449 | 40 | 1 | 41 | 0.363 (0.034) |
| Pelvis BMD | 31873 | 48 | 1 | 49 | 0.351 (0.036) |
| Rib BMD | 31873 | 22 | 0 | 22 | 0.298 (0.030) |
| Spine BMD | 31986 | 37 | 0 | 37 | 0.353 (0.034) |
| Total BMD | 31986 | 61 | 0 | 61 | 0.379 (0.039) |
| Trunk BMD | 31873 | 45 | 1 | 46 | 0.360 (0.036) |
| **All BMD-related phenotypes** | \ | 476 (232 unique) | 11 (8 unique) | 487 (240 unique) | \ |
| Fracture | 352,791 | 8 | 0 | 8 | 0.048 (0.007) |
|  | (N=35,192 for cases;<br>N=317,599 for controls) |  |  |  |  |
| **Acorss phenotypes** | \ | 87 | 5 | 92 | \ |

### Five loci identified for DXA-derived BMD traits

Although previous GWASs have reported hundreds of loci, we still identified five loci for six BMD traits that were not reported previously (**Figure 1B** and **Figure 2 A-E**). Among these loci, the most pleiotropic locus resided between *ABCA1* and *SLC44A1* genes on chromosome 9 (**Figure 1B, Figure 2A**, and **Supplementary Table 4**). SNPs (rs1039406 and rs746100) around this locus were genome-wide significantly associated with five BMD sites, including the lumbar spine, femur neck, femur total, pelvis, and trunk (**Figure 1B, Figure 2A**, and **Supplementary Table 1**). The eQTL data from whole blood tissue revealed that SNP rs746100 was also associated with the gene expression of *ABCA1* (*P*=2.68×10^−5^) in artery tibial tissue, based on the GTEx consortium (**Supplementary Table 5** and **Supplementary Figure 14**), and the genetically predicted higher *ABCA1* gene expression in whole blood tissue was associated with higher BMD (**Figure 2A**). The second promising locus resided between *SWAP70* and *WEE1* genes on chromosome 11 with leading SNP rs10840273, showing a genome-wide significant association for leg BMD (*P*-value=4.52×10^−9^) (**Figure 2B Supplementary Table 1** and **Supplementary Table 4**). Whole blood eQTL data from eQTLGen identified that rs10840273 was associated with the *SWAP70* gene expression (*P*=2.42×10^−39^) (**Supplementary Table 5** and **Supplementary Figure 15**). Mesenchymal stem cell Hi-C data also detected a direct interaction of the associated region with the *SWAP70* gene (FDR-corrected *P*-value_interaction_=2.74×10^−109^) (**Supplementary Table 6**). Furthermore, this lead SNP showed a genome-wide significant association with circulating SWAP70 (*P*-value=6.94×10^−81^). The MR results revealed that genetically predicted higher *SWAP70* gene expression and higher circulating SWAP70 protein in whole blood were significantly associated with increased leg BMD (**Figure 2B**).

Another locus surrounding rs12916774 on chromosome 15 was associated with femur neck and femur total BMD (**Figure 2C** and **Supplementary Figure 16**). Both eQTL data and Mesenchymal stem cell Hi-C data consistently supported the *CHSY1* as a plausible candidate gene (*P*=2.15×10^−53^ for *CHSY1* eQTL in whole blood tissue from eQTLGen; FDR-corrected *P*_interaction_=1.72×10^−49^ for Hi-C data) (**Supplementary Table 5** and **Supplementary Table 6**). The fourth locus (lead SNP: rs6013897) was an intergenic region of *CYP24A1* and *BCAS1* (**Figure 2D, Supplementary Table 4** and **Supplementary Figure 17**). The Mesenchymal stem cell Hi-C data detected a direct interaction of the associated region with the *CYP24A1* gene (FDR-corrected *P*_interaction_=8.04×10^−78^) (**Supplementary Table 6**). We further prioritized *PAX1* as a potential candidate gene for rs927059, which is a lead SNP for femur neck BMD (*P*-value=1.87×10^−8^) (**Figure 2E, Supplementary Table 4** and **Supplementary Figure 18**). The positional and eQTL annotation results consistently supported the *PAX1* as a candidate gene for rs927059 (*P*-value=4.40×10^−6^ for *PAX1* eQTL in muscle skeletal tissue from GTEx) (**Supplementary Table 4** and **Supplementary Table 5**). In summary, using multi-omics datasets, we prioritized 5 potential candidate genes (i.e., *ABCA1*, *CHSY1*, *CYP24A1*, *SWAP70,* and *PAX1*) to 5 novel loci (**Figure 1B, Figure 2 A-E**, and **Supplementary Table 4-6**). The annotation results for other known loci have also been shown in **Supplementary Table 4-7**.

### Polygenic risk score demonstrated marginal improvement in fracture prediction

Based on effect size derived from GWASs 11 DXA-BMD traits, heel BMD, and fracture in training datasets, we selected SNPs that could achieve the best predictive PRSs for fracture in the validation dataset (**Figure 3A**), resulting in 29 (for rib BMD)-79292 (for head BMD) selected SNPs for different trait (**Supplementary Table 8**). After obtaining SNPs and the effect size for each trait, we calculated the corresponding PRS for each participant in the test dataset (**Figure 3A**). The metaPRS was generated by integrating these 13 individual PRSs using stepwise Cox regression in the validation cohort dataset (**Figure 3A**), with estimates for each single PRS contained in the best-performing model (**Supplementary Table 9**). The association of metaPRS with fracture incidence was largely independent of the traditional risk factors (**Supplementary Table 10**). As illustrated in **Figure 3B**, most individual PRSs showed significant associations with fracture risk in the test cohort dataset (*P*_< 0.05) after adjusting for age, sex, obesity, smoking, alcohol, glucocorticoid medicine use, BMD, and population stratification. However, these PRSs exhibited similar effect estimates for fracture risk, with the metaPRS displaying the most prominent association [HR: 1.134, 95% confidence interval (CI) 1.098-1.172, *P*_=_4.15×10^−14^] (**Figure 3B**). Furthermore, we observed a more marked gradient of fracture risk across quintiles of metaPRS (HR=1.364, 95% CI=1.243-1.498) than fracture PRS (HR=1.177, 95% CI=1.077-1.287) in the top quartile vs. the bottom quartile (**Supplementary Table 11**).

**Figure 3.**
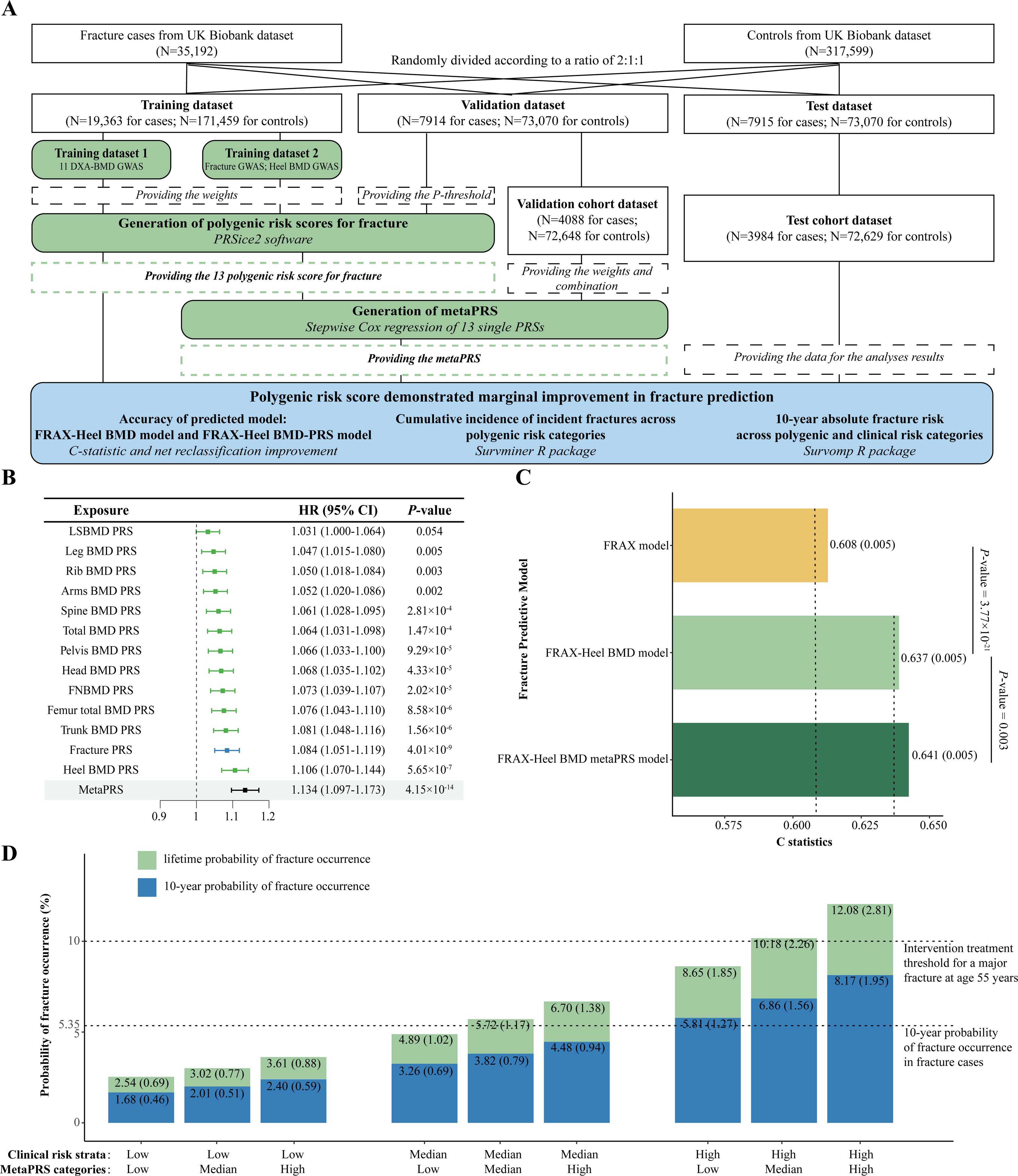
PRS demonstrated marginal improvement in fracture prediction. (A) study design; (B) Forest plot of the association of each PRS with fracture risk, adjusting for age, sex, obesity, smoking, alcohol, glucocorticoids medicine use, heel BMD, and population stratification (the first five principal components); (C) The fracture predictive results of three models. The FRAX model included clinical risk factors from FRAX tools [i.e., sex, age, obesity, current smoking, current alcohol consumption, and glucocorticoids medicine use]. For the FRAX-heel BMD model, heel BMD was integrated with on the FRAX model. For the FRAX-heel BMD metaPRA model, metaPRS was integrated into the FRAX-heel BMD model; (D) The lifetime and 10-year probability of fracture occurrence across metaPRS categories within each clinical risk strata. Abbreviations: PRS, polygenic risk score

By including only clinical factors such as age, sex, BMI, smoking, alcohol use, and glucocorticoid use (the FRAX model), we observed limited predictive performance of this model (C-statistic=0.608, sd=0.005) (**Figure 3C** and **Supplementary Table 12**). We found that adding BMD to the FRAX model increased the C-statistic from 0.608 to 0.637 (difference, 4.77%, *P*=3.77×10^−21^) (**Figure 3C** and **Supplementary Table 12**). However, the addition of various PRS to the FRAX-BMD model did not substantially improve the C-statistic (**Figure 3C** and **Supplementary Table 12**). Incorporating metaPRS into the FRAX-BMD model resulted in the highest C-statistic (C-statistics = 0.641), with a C-statistic change of 0.63% (*P*=0.003), compared with the FRAX-BMD model (**Figure 3C** and **Supplementary Table 12**). By utilizing the optimal cutoff point from the FRAX-BMD metaPRS model as the threshold, the combination of metaPRS and FRAX-BMD model yielded a moderate improvement in net reclassification improvement (NRI=1.66%, 95% CI 0.7%-2.62%; the continuous NRI: 9.15%, 95% CI 6.39%-11.91%) (**Supplementary Table 13**).

We further assessed how the interplay of the metaPRS and clinical risk factors impact the fracture risk. Firstly, we found that the cumulative incidence for fracture events was 4.63% for individuals with low polygenic risk (bottom quintiles of the metaPRS) and 7.58% among those with high polygenic risk (top quintiles of the metaPRS), suggesting that metaPRS could stratify individuals into different trajectories of fracture risk (**Supplementary Figure 19**). Similar results were observed in both sexes, with women having higher HR (**Supplementary Table 11**) and higher cumulative risk (**Supplementary Figure 20** and **Supplementary Figure 21**). Although we observed significant gradients in the 10-year probability of fracture occurrence across metaPRS categories within each clinical risk strata (**Figure 3D**), the clinical risk factors played more important role in the stratification. For example, among participants with low clinical risk, the 10-year probability of fracture occurrence for those with high genetic risk (2.40% sd=0.59%) was yet lower than the participants with median clinical risk but low genetic risk (3.26%, sd=0.69%) (**Figure 3D**). And the 10-year probability of fracture of the participants at high clinical risk with low genetic risk (5.81%, sd=1.27%) had already exceeded the 10-year probability in fracture cases only (5.35%, sd=2.63%) (**Figure 3D**). The lifetime risk of incident fracture was higher in each stratum than the 10-year probability (**Figure 3D**). Participants at high clinical risk with median/high genetic risk demonstrated lifetime probabilities of 10.18% and 12.08%, surpassing the intervention treatment threshold of 10% for a major fracture at age 55 years when treatment should be recommended ^19^(**Figure 3D**).

### The shared genetic architecture of head BMD and intracranial aneurysm

We further estimated the shared genetic architecture of DXA-BMD at 11 sites with other 13 common chronic diseases, including neurodegenerative diseases, cardiovascular diseases and autoimmune diseases (**Supplementary Table 14**). First of all, we tested the pair-wise correlation between the BMD traits. It is suggested that there were the weakest correlations for head BMD with other BMD traits in both phenotypic and genetic correlation analyses, although all pairs exhibited statistically significant phenotypic correlation (**Figure 4A**). In the 143 BMD-disease pairs (11 BMD traits × 13 diseases), we only observed a statistically significant inverse genetic correlation of head BMD with intracranial aneurysm (IA) (r_g_=-0.188, se=0.055, FDR-corrected *P*=0.0096), while the genetic correlations with other 12 common chronic diseases were not significant (FDR-corrected *P*>0.05) (**Figure 4B** and **Supplementary Table 14**). Furthermore, no significant genetic correlation was observed for the remaining 10 DXA-BMD traits with IA (FDR-corrected *P*>0.05) (**Figure 4B** and **Supplementary Table 14**). Compared to the specificity of observed genetic correlation, there was a similar MiXeR estimated polygenic overlap between head BMD and IA. 29.36% (N=114, SD=15) of the 390 head-BMD influencing variants were also predicted to influence IA (**Figure 4C** and **Supplementary Table 15**). By employing the conjFDR method, we identified four genomic loci jointly associated with head BMD and IA (**Figure 4D, Figure 4E**, and **Supplementary Table 16**). Intriguingly, 3 of the 4 lead SNPs (rs72560793, rs10958404, rs11187838) had the opposite effect direction, consistent with the moderate inverse genetic correlation between head BMD and IA (**Figure 4E**, and **Supplementary Table 16**). Notably, two of the four loci demonstrated strong evidence of colocalization (H4>0.5), suggesting the presence of shared causal variants between head BMD and IA (H4: 0.809 for rs10832558 within *SOX6*; H4: 0.581 for rs11187838 within *PLCE1*) (**Supplementary Table 17**). Genes mapped to these shared loci were enriched for biological processes and cellular components related to the skeletal systems (e.g., positive regulation of chondrocyte differentiation) and vascular smooth muscle (i.e., regulation of Ras protein signal transduction) (**Supplementary Table 18**).

**Figure 4.**
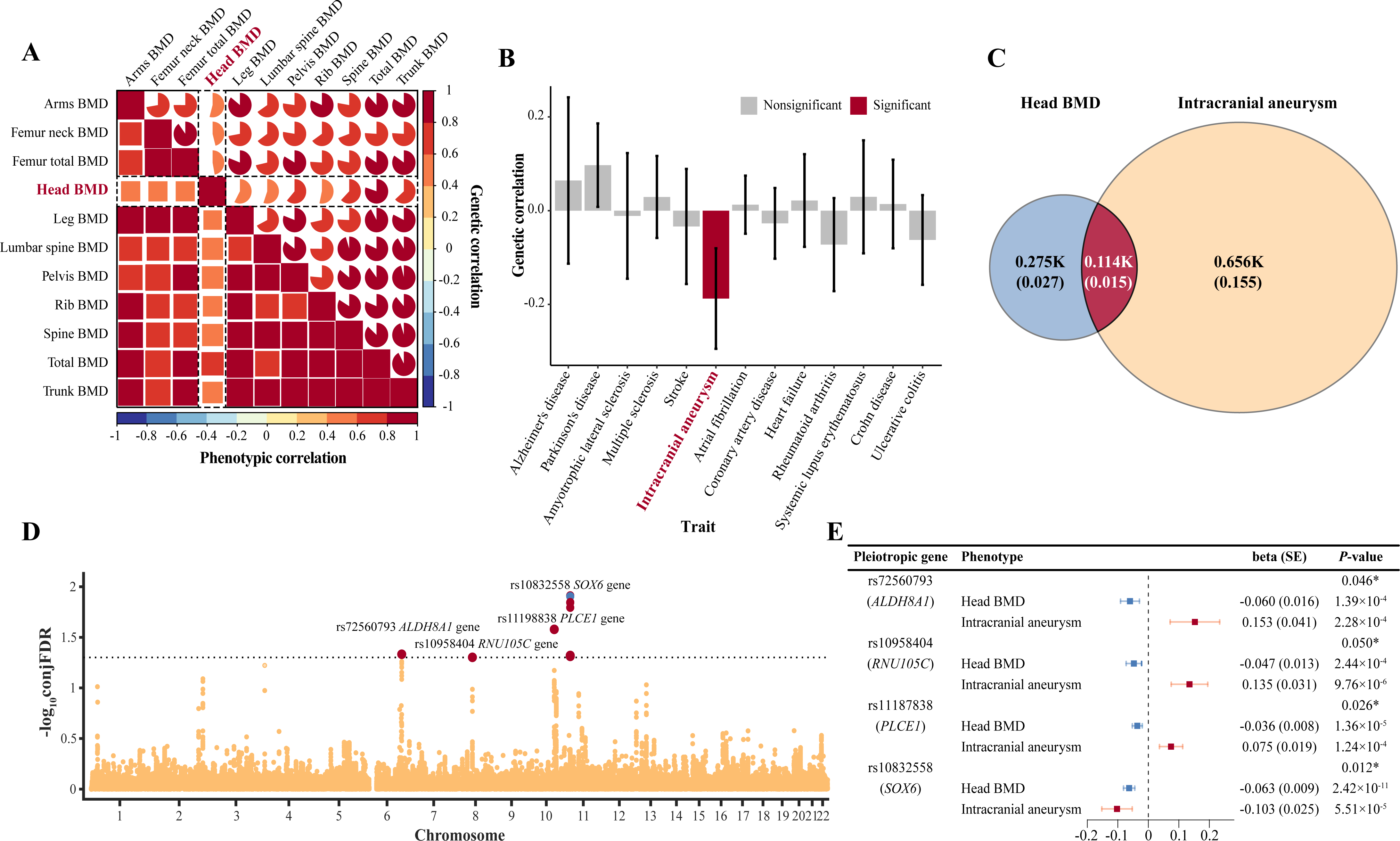
The shared genetic architecture of head BMD and intracranial aneurysm. (A) heatmap of genetic and phenotypic correlation between 11 DXA-derived BMD; (B) the genetic correlations of head BMD with 13 common chronic diseases; (C) Venn diagrams of shared variants between head BMD and intracranial aneurysm, and unique variants per trait; (D) Shared loci between head BMD and intracranial aneurysm. Common genetic variants jointly associated with head BMD and intracranial aneurysm at conjFDR < 0.05 were highlighted in red. (E) Forest plot of the genetic association estimates of four joint-associated variants with head BMD and intracranial aneurysm. Abbreviations: BMD, bone mineral density.

### Prioritization of drug targets

Subsequently, by integrating the druggable genome, gene expression, and GWAS datasets, we aimed to identify the genetically supported potential therapeutic targets for osteoporosis, emulating exposure to corresponding medications. Utilizing drug target information from the ChEMBL database (release 29), we included a total of 3,329 druggable genes for subsequent analyses. Next, we employed eQTL data from muscle (including 791 druggable genes), artery tibial (917 druggable genes), and whole-blood tissue (845 druggable genes from GTEx; 2104 druggable genes from eQTLGen) to test the association with BMD through mendelian randomization approach. We observed statistically significant associations between genetically predicted expression of 15 genes and DXA-BMD (FDR-corrected *P* < 0.05) (**Supplementary Table 19**). Among these, genetically predicted expressions of 4 genes (*CCR1, ESR1, NCOR1* and *SREBF1*) were associated with at least two DXA-BMD traits with consistent direction, providing robust MR evidence for the genes (**Figure 5A, 5B** and **Supplementary Table 19**). For these four genes, genetically predicted *ESR1* gene expression showed negative associations with 9 DXA-BMD traits (**Figure 5A** and **5B**). There were positive associations of genetically predicted *NCOR1* gene expression with head BMD and total BMD, while negative associations were found for *SREBF1* and *CCR1* gene expressions (**Figure 5A** and **5B**). To assess whether the genetic association between these gene expressions and phenotypes shared the same causal variant, we conducted colocalization analyses of the genes with DXA-BMD traits. We discovered that eQTLs in whole blood tissue for 3 genes (i.e., *SREBF1*, *NCOR1* and *CCR1*) colocalized with DXA-BMD loci (H4>0.5), reinforcing the evidence for these genes as drug targets for DXA-BMD (**Figure 5C** and **Supplementary Table 20**). Considering the observed negative association between *SEEBF1* and *CCR1* gene expression and BMD (**Supplementary Table 19**), there were relevant inhibitors/antagonists that have been approved or under investigation that present possible repurposing opportunities for osteoporosis treatment. Specifically, SEBF1 could be targeted using Doconexent (inhibitor) and Omega-3 fatty acids (inhibitor), while CCR1 could be targeted using CCX354-C (antagonist) (**Figure 5D**).

**Figure 5.**
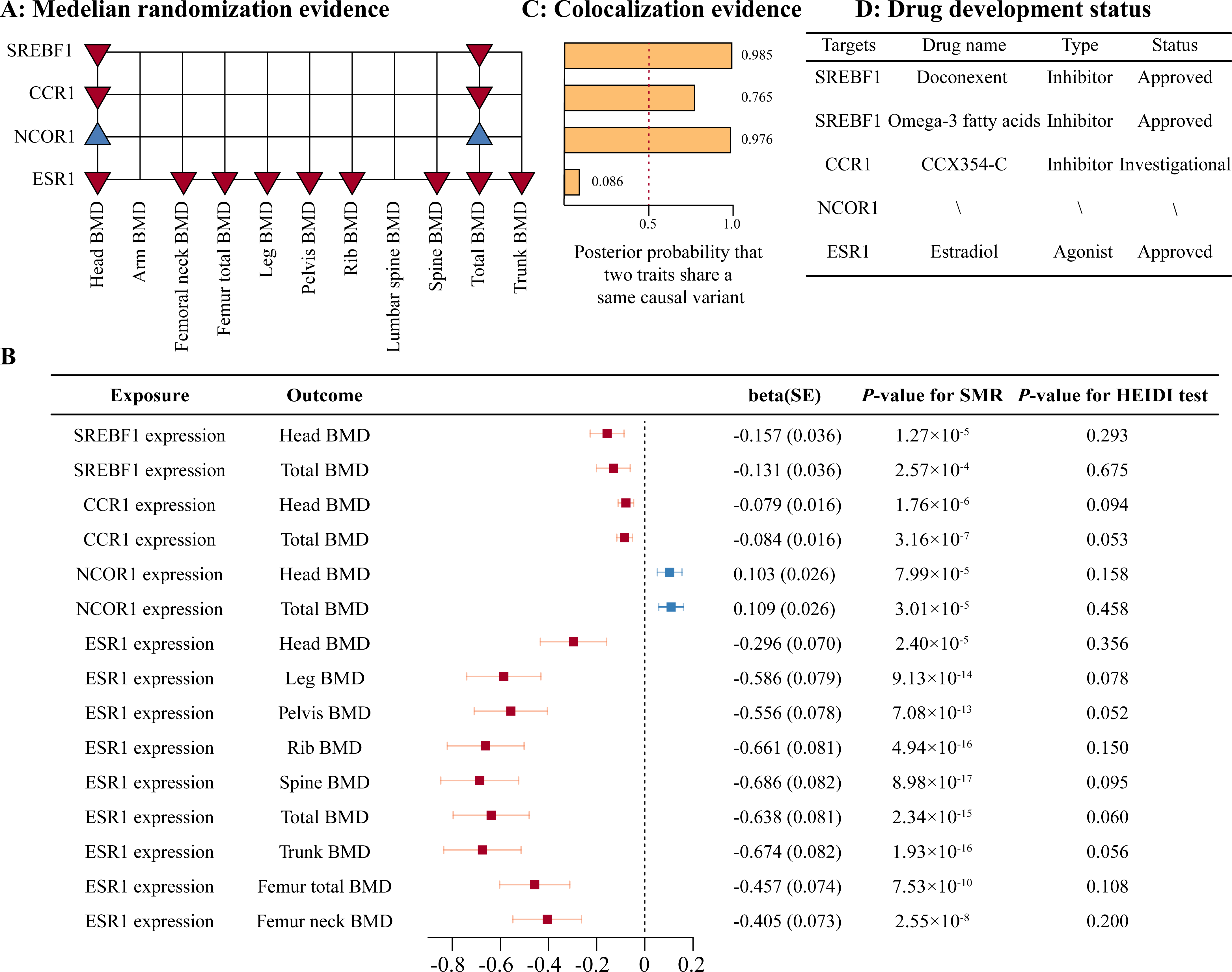
Prioritization of drug targets. (A) the result of four genes with mendelian randomization evidence. The red downward triangle indicates that genetically predicted expression level of this gene is negatively correlated with BMD, while the blue upward triangle indicates a positive association for BMD; (B) Forest plot of the association of genetically predicted *SREBF1*, *CCR1*, *NCOR1* and *ESR1* gene expression with BMD, based on summary-based mendelian randomization analyses; (C) colocalization results of GWAS and eQTL within *SREBF1*, *CCR1*, *NCOR1* and *ESR1* gene regions; (D) the drug development status of *SREBF1*, *CCR1*, *NCOR1* and *ESR1* genes. Abbreviations: BMD, bone mineral density; eQTL, expression quantitative trait locus; GWAS, genome-wide association study.

## Discussion

In this study, we first conducted the large-scale GWASs of DXA-BMD at 11 skeletal sites, and identified 91 unique and independent loci associated with at least one phenotype, including five previously unreported BMD loci for six BMD traits (i.e., *ABCA1*, *CHSY1*, *CYP24A1*, *SWAP70* and *PAX1*). These novel loci exhibited evidence of association in both male and female, which could serve as indepdent replication. Additionally, the incorporation of multiple PRSs (metaPRS) with the clinical risk factors (i.e., the FRAX model) exhibited the highest predictive performance, however, the improvement was marginal in fracture prediction. Although the metaPRS could stratify individuals into different trajectories of fracture risk, the clinical risk factors played a more important role in the stratification. We further estimated the shared genetic architecture of DXA-BMD at 11 sites with other common chronic diseases, including neurodegenerative diseases, cardiovascular diseases and autoimmune diseases, and we only uncovered genetic correlation and shared polygenicity between head BMD and intracranial aneurysm. And the gene *PLCE1* might play important roles in the shared polygenicity. Finally, by integrating the gene expression and GWAS datasets, we prioritized drug targets (e.g. *ESR1*, *SREBF1*, *CCR1* and *NCOR1*) within the druggable genomic genes along with their respective inhibitors/antagonists.

Although previous GWAS have identified hundreds of association signals ^1,2^, we considered reporting five loci in this study when the associated SNPs improved at least two orders of magnitude of significance compared to the most significant SNPs within the region (position-of-reported-SNP±250 kb) in any of the previous BMD GWASs. For example, in our study, the locus (rs746100) near *ABCA1* was associated with five BMD traits, including the lumbar spine, femur neck, total femur, pelvis, and trunk with the smallest *P*-value at 1.64×10^−9^. By looking back at the meta-analysis of GWAS in a relatively large sample size (N=∼30,000), the SNP rs1831554 within this locus had a marginal significance for femur neck (*P*=9.94×10^−5^) and lumbar spine (*P*=1.41×10^−4^) BMD ^8^. The pair-wise LD of the two lead SNPs was 0.0005. In our study, we used the individual-level genotype data within ∼30,000 samples, the sample size was as large as the GWAS meta-anlaysis of summary statistic data ^8^, but the association significance improved greatly. It is suggested that the association analysis performed in individual-level genotype data could enable a more comprehensive power to control various factors, such as population structure, covariates, and phenotype definitions ^20^. Another example was the locus near the *CHSY1* gene, this locus showed marginal significance (*P*-value=2.30×10^−5^ for rs3784491) in the largest-scale GWAS to date for QUS-derived heel BMD ^10^, the sample size was more than ten times compared to our study, however, the SNP rs12916774, with very low LD with rs3784491 (LD r^2^=0.005), was found to be genome-wide significantly associated with femur neck BMD in our study (*P*=2.14×10^−9^). It should be noted that QUS-derived BMD primarily reflected the bone mass at the heel calcaneus and exhibited limited correlation (0.5∼0.65) with DXA-derived BMD at the spine and hip ^21^. Additionally, we confirmed the *ZIC1*/*ZIC4* locus for head BMD (*P*=2.19×10^−8^) which was reported in a very recent GWAS meta-analysis ^22^.

One of the potential applications of genetic data is disease prediction ^1^. Lu et al calculated the genetically predicted speed of sound (SOS, measured by quantitative ultrasound at the heel) for individuals in the UK Biobank and assessed the predictive performance of this score ^11^. In this study, we used three independent datasets and generated PRSs for the DXA-derived BMD at multiple skeleton sites. Our results indicated that PRSs had robust associations with incident fracture, even after adjusting for the related clinical risk factors such as age, sex, obesity, smoking, alcohol, glucocorticoid use and BMD, suggesting the independent contribution to the susceptibility of fracture. We further built metaPRS by combining multiple PRSs for DXA-BMD, heel BMD, and fracture to evaluate the potential of PRSs on fracture prediction. As expected, the metaPRS showed a larger effect size on fracture risk than fracture PRS. This improvement could be attributed to that the genetic component of this metaPRS captured the majority of the genetic basis of fracture. At baseline, we included the FRAX factors ^23^ in the prediction model, and only limited predictive performance was observed just as before ^24^. We observed an increased C-statistic when incorporating BMD into the FRAX model. However, the addition of various PRS to the FRAX-BMD model did not substantially improve the C-statistic, suggesting that the predictive performance of PRS did not perform as well as BMD measurement itself. Additionally, the probability of fracture occurrence for those with low clinical risk and high genetic risk was yet lower than the participants with median clinical risk but low genetic risk, suggesting that the clinical risk factors played a more important role in the stratification. Lu et al suggested that the predictive performance of genetically determind SOS surpassed single clinical risk factor such as smoking, corticosteroids use and falls etc ^11^, but they did not test the combination of the these risk factors. Consistently, the predictive performance of PRS would not outperform BMD ^11^.

Clinically, intracranial aneurysm (IA) is characterized by a bulge or distention of an artery in the brain due to weakness and inelasticity of the vessel wall ^25^. The disruption of the extracellular matrix (ECM) has been proposed as a contributing factor in the pathophysiology of IA ^26^. The ECM is a also salient feature of bone tissue. Bone ECM, containing minerals deposited on highly crosslinked collagen fibrils, dynamically interacts with osteoblasts and osteoclasts to regulate the process of bone regeneration ^27^. Given the shared histological basis of bone and vessel, the genetic correlation analysis in this study suggested that higher head BMD would associated with a lower risk of IA. This genetic association was supported by an epidemiological study that the IA risk was increased in patients with BMD in middle and lower tertiles compared with patients with BMD in higher tertile ^28^. Further, with conditional false discovery rate approach ^29^, we identified four shared signals, emphasizing the pleiotropic effect underlying BMD and IA. Two of them demonstrated evidence of colocalization (rs10832558 near *SOX6* and rs11187838 near *PLCE1*). The SNP rs10832558 was at the same effect direction for head BMD and IA, which was not consistent with the inverse genetic correlation. Here, we highlighted the variant rs11187838 shared by BMD and IA with opposite effect direction, which had not been detected by both previous single-trait analyses. This SNP was mapped to the *PLCE1* gene, encoding the enzyme phospholipase C epsilon-1. This enzyme could stimulate the Ras and mitogen-activated protein kinase (MAPK) signaling pathway through the regulation of heterotrimeric G protein Galpha ^30^. Ras signaling stimulated the proliferation of immature osteoprogenitor cells to increase the number of osteoblastic descendants in a cell-autonomous fashion ^31^. Additionally, the activation of Ras/MAPK signals could stimulate the migration and proliferation of vascular smooth muscle cells through fibronectin ^32^. These synthetic vascular smooth muscle cells could secrete large amounts of ECM components, including collagen, elastin, and matrix metalloproteinase, causing vascular ECM remodeling ^33^. All these results suggested that *PLCE1* might play important roles in the shared polygenicity between BMD and IA. Finally, we did not observe genetic correlations between BMD and other diseases in our study. As the global genetic correlation represented the average of genome-wide shared association, the nonsignificant global correlation might be due to opposing directions at different genomic regions^34^.

Several pharmacological agents were available to osteoporosis patients, either by reducing bone resorption such as bisphosphonate and denosumab, or by stimulating bone formation such as teriparatide and abaloparatide ^35^. The fruitful GWAS discoveries in the bone field have proven useful in identifying compounds suitable for drug repurposing ^36^. One possible approach is to use genetic variants associated with the expression level of a gene encoding druggable human protein to proxy the lifelong exposure to a medication targeting corresponding gene production ^37,38^. This Mendelian randomization (MR) approach could mimic a randomized controlled trial to cost-effectively predict the treatment response of a drug ^37,38^. In this study, by using GWAS data and eQTL data, we prioritized several drug targets for osteoporosis such as ESR1 and SREBF1, etc. The estrogen hormone therapy, targeting ESR1 protein, was an old-fashioned treatment for osteoporosis and was rarely used nowadays because of the adverse side effects such as cardiovascular conditions and cancer ^39^. The SREBF1 we would highlight here was the target of Doconexent (a high-docosahexaenoic acid supplement) and Omega-3 fatty acids. Daily marine omega_3 supplementation had been widely recommended in the prevention of adverse coronary events ^40,41^. However, the effect of this kind of fatty acid on bone health is controversial. For example, a meta-analysis of 23 randomized controlled trials did not show any significant effect of n-3 PUFA supplementation on BMD at any body’s part ^42^. Nevertheless, when subgroup analyses were performed, it was observed that the impact of n-3 PUFA supplementation on BMD varied across different regions ^42^. Specifically, individuals from Eastern countries exhibited higher BMD at the lumbar spine and femoral neck following n-3 PUFA supplementation, in comparison to individuals from Western countries ^42^. However, another systematic review and meta-analysis of randomized controlled trials suggested that n-3 PUFAs might have a beneficial effect on bone health, especially for postmenopausal women ^43^. In our study, we revealed a negative association between *SREBF1* gene expression and BMD. Previous studies suggested that the supplement of omega-3 polyunsaturated fatty acid negatively regulated SREBF1 ^44,45^. And decreased expression of the *SREBF1* gene could inhibit osteoclast formation and bone resorption activity by decreasing NF-κB signaling ^46^. Therefore, we hypothesized that the n-3 PUFA supplementation might be effective for the prevention of osteoporosis. For CCR1 antagonist, BMS-817399 failed in Phase 2, double-blind, placebo-controlled clinical trial ^47^, while another CCR1 antagonist (CCX354-C) has shown a good safety and tolerability profile and evidence of clinical activity in rheumatoid arthritis in Phase II trials (NCT01242917) ^48^. Previous animal study have shown that the activation of CCR1 leads to the formation of osteolytic lesions through the regulation of CCL3 ^49^.

In conclusion, we conducted large-scale GWASs of DXA-derived BMD traits and identified novel signals that will likely provide new insights into the biological mechanism of osteoporosis. We demonstrated that although PRSs were independently associated with fracture risk, the predictive performance improved marginally compared to the clinical risk factors. Additionally, we uncovered a genetic correlation between head BMD and IA, and the joint associated genes such as *PLCE1* might play important roles in the shared genetic basis. Finally, the prioritization of genetically-supported targets implied the potential repurposing drugs (for example the n-3 PUFA supplements targeting SREBF1) for the prevention of osteoporosis.

## Materials and methods

### Source of the phenotypes and quality control of the genotype

As we used before ^50–52^, the individual-level data from the UK biobank (Application 41376) was used for discovery analyses. The UK Biobank is a cohort of roughly ∼500,000 participants aged 40-69 years, of which, 487,409 participants were genotyped with the UK Biobank Axiom or UK UKBiLEVE Array, and then imputed by the 1000 Genomes Project (Phase 3) reference panel ^53,54^. Ethics approval for the UK Biobank research was obtained from the North West Multicentre Research Ethical Committee, and all participants provided informed consent (original ethics committee approval number: 21/NW/0157). In this study, we extracted BMD traits measured by dual-energy X-ray (DXA) from 11 anatomical sites (i.e., arm, total femur, femoral neck, head, legs, lumbar spine, pelvis, ribs, spine, trunk, and total body) and fracture as phenotypes (**Figure 1A** and **Supplementary Table 21**). The fracture cases were defined as participants with the diagnosis of any site of fracture (except fractures with known primary diseases and those with diseases that might affect bone health) (**Supplementary Table 21**). To minimize the population stratification bias, we further excluded participants who were not of European ancestry (**Supplementary Table 21**), and those who had a kinship with any participants. For quality control of genotype data, the variants were excluded if the minor allele frequency (MAF) < 0.01, imputation info score < 0.3, missing genotype rates > 0.05, and *P*-value for Hardy–Weinberg equilibrium test < 1 × 10^−6^. After the quality control, a total of 5,996,792 imputed variants and around ∼30,000 participations (**Figure 1A** and **Table 1**) remained for BMD GWAS analysis, as well as 352,791 participants (N=35,192 for cases; N=317,599 for controls) for fracture GWAS (**Figure 1A** and **Table 1**).

### Genetic association analysis of BMD and fracture

To identify the genetic variants associated with BMD at a genome-wide significant level (*P* ≤ 5×10^−8^), we conducted the GWAS analyses on BMD traits at 11 skeletal sites. For BMD at each site, the values (g/cm^2^) were stratified by sex, and then adjusted for age, age^2^, weight, menopause status (only for females), and first 5 principal components using linear regression. The standardized residuals (mean=0 and sd=1) in males and females (i.e., standardized BMD) were used in the GWAS analyses. The associations between genetic variants with phenotypes (i.e., standardized-BMD at 11 skeletal sites) were analyzed using the PLINK software (http://www.cog-genomics.org/plink2/). We then combined the summary statistics of the two sexes by an inverse variance weighted fixed effects meta-analysis, using the METAL software ^55^. We also analyzed the association between genetic variants and fracture risk, adjusting for sex, age, weight, and the first 5 principal components using the PLINK software. The lead SNP of novel loci with *P*-value from sex-stratified GWASs less than 0.05 were considered to be replicated.

### Identification of statistical independence and novel loci

The conditional independent signals for each BMD trait (between-sex meta-analysis) were defined using the conditional and joint (COJO; gcta --cojo-slct) analysis^56^. 10,000 randomly selected unrelated white British individuals from the UK Biobank were used as linkage disequilibrium (LD) references. The conditional independent SNV for each signal was defined as the SNV with both *P*-value for original GWAS and *P*-value for COJO joint analyses less than 5 ×10^−8^. Among these independent signals, the association was classified into the “novel” signal if all SNPs within one signal (conditional independent SNV ± 250 kb) have not been reported to be significantly associated with BMD (*P*<1×10^−6^) in previous BMD GWASs ^8,10,57^. Across 11 BMD traits, the identified conditional independent significant SNVs were merged into one locus if they were closely located to each other (<500 kb), leaving the SNP with the smallest *P*-value as the lead SNP. The pleiotropic genomic locus was defined as a genomic locus containing multiple conditional independent signals for different BMD traits.

### Variant annotation

We then used the ANNOVAR software ^58^, and Functional Mapping and Annotation of Genome-wide Association Studies (FUMA, https://fuma.ctglab.nl/)^59^, as well as Online Mendelian Inheritance in Man database (OMIM, http://omim.org/) ^60^, to obtain functional annotation for conditional independent significant SNVs. Specifically, these SNVs were first physically annotated using ANNOVAR software. Based on the FUMA website, we further obtained the eQTL and chromatin interaction annotation results. We selected eQTL datasets from eQTLGen Consortium and five tissue types (i.e., artery tibial, whole blood, and muscle-skeletal) based on the Genotype-Tissue Expression project (GTEx v8), and long-range interactions (Hi-C) dataset from GSE87112 (Mesenchymal stem cell). Additionally, we performed the gene map search in OMIM dataset using ‘(OSTEOPOROSIS OR “bone fragility” OR “fragile bones” OR “bone mineral density”)’ to obtain gene list for BMD phenotype. For each physical annotated genes, we collected corresponding evidence codes from the above datasets (p for physical annotation; e for eQTL annotation; h for HiC annotation; o for OMIM results).

### Integrating polygenic risk score with clinical risk score for risk stratification of fracture

#### Training, validation, and test datasets

We evaluated the potential clinical utility of polygenic risk scores (PRSs) for fracture incidence combined with traditional clinical risk factors. Here, two training datasets were set in the analyses for DXA-derived BMD (training dataset 1) and heel BMD/fracture (training dataset 2), respectively (**Figure 3A**). The training dataset 1 was derived from the aforementioned DXA-derived BMD GWAS. Additionally, all fracture cases (N=35,192) and controls (N=317,599) from the UK biobank were randomly divided into three distinct datasets: training dataset 2 (N=171,459 for controls, N=19,363 for cases), validation (N=73,070 for controls, N=7914 for cases), and test (N=73,070 for controls, N=7915 for cases). These divisions were conducted according to a ratio of 2:1:1. Following this, both heel BMD GWAS and fracture GWAS analyses were performed utilizing the aforementioned GWAS pipeline in training dataset 2 (**Figure 3A**).

#### Generation of polygenic risk scores (PRSs)

Based on GWAS summary statistics from 13 traits (i.e., 11 DXA-derived BMD traits, heel BMD and fracture) in training datasets, we then used the PRSice 2 software ^61^ to implement the clumping and threshold approach for developing PRSs for fracture in the validation dataset (**Figure 3A**). The best predictive PRSs were assessed for transferability and predictivity through the *P*-values and Nagelkerke R^2^ in logistic model implemented in PRSice 2 software ^61^, which corrected for age, sex, weight and population stratification (first five principal components). After obtaining the *P*-values threshold for the best predictive PRS from the validation dataset, we calculated the corresponding PRS for each participant in the test dataset (**Figure 3A**).

#### Generation of metaPRS

To generate a combined PRS (i.e., metaPRS), we first removed the 4,248 participants with fracture history at the baseline to generate a validation cohort dataset (N=72,648 controls; N=4,088 cases) (**Figure 3A**). Based on this validation cohort dataset, we included all 13 PRSs and conducted stepwise Cox regression in the validation cohort dataset, which could automatically select a reduced number of predictor variables for building the best-performing Cox regression model. Accordingly, we computed the metaPRS by summation of single PRS (which were contained in the best-performing model), weighted by beta value from stepwise Cox regression.

#### Prediction fracture risk

In this analysis, based on test datasets, we further removed the participants with fracture history at the baseline, leaving 76,613 participants as the test cohort datasets for fracture (N=72,629 controls; N=3,984 cases) (**Figure 3A**). We first generated a basic FRAX-BMD model including clinical risk factors from FRAX tools [i.e., sex (categorical: male and female), age (continuous: years), obesity (categorical: 1^st^, BMI ≤20; 2^nd^, 20<BMI≤25; 3^rd^, 25<BMI≤30; 4^th^, 30<BMI≤35; 5^th^, 35<BMI≤40;6^th^, 40<BMI≤ 45; 7^th^, BMI>45), current smoking (categorical: yes and no), current alcohol consumption (categorical: yes and no), and glucocorticoids medicine use (categorical: yes and no)] and heel BMD (**Supplementary Table 21**). Using Cox regression for fracture, we obtained the predicted values based on the basic FRAX-BMD model in the test dataset. We then employed C-statistic as a quantitative measure to evaluate the accuracy of the basic FRAX-BMD model using these predicted values in the same dataset. Additionally, we quantify the variations in discriminative power when integrating various PRSs into the basic FRAX-heel BMD model (FRAX-heel BMD PRS model). Specifically, for each type of PRS (i.e., heel BMD, 11 DXA-BMD, fracture and metaPRS), we performed a multiple Cox regression for fracture adjusting for age, sex, obesity, smoking, alcohol, glucocorticoids medicine use, heel BMD, and population stratification (the first five principal components). Based on these predicted values, C-statistics and net reclassification improvement (NRI) were used to estimate the improvement in discrimination and reclassification after adding the various PRSs to the basic FRAX-BMD model. The C-statistics change was calculated by (C-statistics_FRAX-heel BMD PRS model_-C-statistics_FRAX-heel BMD model_)/(C-statistics_FRAX-heel BMD model_-0.5)*100%. The difference of C-statistics from various FRAX-heel BMD PRS models were estimated based on student t-test using *cindex.comp* function from “survomp” R package. The optimal cutoff point, which was obtained from the FRAX-BMD metaPRS model, was utilized to calculate NRI.

Additionally, to visualize the cumulative incidence of incident fractures across polygenic risk categories (i.e., low (bottom quartile), intermediate (the second to the third quartile), and high (top quartile) polygenic risk categories according to the quintiles of the metaPRS), we employed the “survminer” R package in the test cohorts consisting of time-to-fracture information and corresponding fracture events as well as polygenic risk categories. We also utilized the ‘cuminc’ function to calculate the cumulative incidence curves. Based on the *survfit* function from “survomp” R, we estimated the 10-year absolute fracture risk, and then assessed the interplay of metaPRS and the clinical risk score (from the basic FRAX-heel BMD model) in impacting the risk of fracture.

### Shared genetic basis of BMD and common chronic diseases

#### Genetic correlation and polygenic overlap

In this study, we first estimated the phenotypic correlation between 11 DXA-derived BMD traits (i.e., arms, femur neck, total femur, head, leg, lumbar spine, pelvis, rib, spine, total body and trunk BMD) using spearman correlation. We then supplied the genetic correlation among them using GCTA software, considering the sample overlap. Additionally, we performed linkage disequilibrium score regression (LDSC) analyses ^62^, based on 1000 Genomes Project European panel, to assess the genome-wide genetic correlation (r_g_) between DXA-BMD and 13 selected common chronic diseases, including neurodegenerative diseases (Alzheimer’s disease, Parkinson’s disease, amyotrophic lateral sclerosis and multiple sclerosis) ^63–66^, cardiovascular diseases (stroke, intracranial aneurysm, atrial fibrillation, coronary artery disease and heart failure) ^67–71^ and autoimmune diseases (rheumatoid arthritis, systemic lupus erythematosus and inflammatory bowel diseases) ^72–74^. For BMD phenotypes with statistically significant genetic correlation, we supplied the bivariate causal mixture model (MiXeR) to quantify the polygenic overlap between BMD and selected chronic diseases beyond genetic correlations ^29^. For a pair of phenotypes, MiXeR estimated the number of trait-influencing SNPs (i.e., SNPs with effects on the disease not inducted by LD) for each trait and the number of shared trait-influencing SNPs based on a bivariate Gaussian mixture model ^29^.

#### Discovery of the shared risk loci

To discover the pleiotropic genetic variants, we performed conditional/conjunctional false discovery rate (condFDR/conjFDR) analysis using genetic summary statistics. We limited our analysis to BMD phenotypes that have evidence to support the shared genetic architecture with common chronic diseases. Based on an empirical Bayesian statistical framework, in the condFDR method, the association between variant and secondary phenotype was used to re-ranks the test statistics and re-calculate the association of this variant with primary phenotype ^75,76^. The conjFDR is determined as the maximum of two condFDR values, which provides a conservative estimate of the posterior probability that a genetic variant showed association with either trait ^75,77^. In this study, the shared genetic variants were defined as variants with conjFDR <0.05. For these identified risk loci with shared effects, we further used the “coloc” R package to determine whether the association signals for DXA-derived BMD and common chronic diseases would co-localize at the shared loci. After extracting genetic association estimates for variants within 250kb of the lead SNP, the probability of H4 that the two traits share one causal variant were calculated. The loci with a probability of H4>0.5 were considered to colocalize ^78^.

### Genetic-driven prioritization of drug targets

The therapeutic target lists were obtained from the ChEMBL database (release 29), which curates the drug information from multiple sources (e.g., United States Adopted Name applications, ClinicalTrials.gov, and FDA Orange Book database) ^79^. Specifically, based on the targets search results, the proteins with values of activity term ≤ 100 and organisms from homo sapiens remained. Accordingly, a total of 3,329 unique druggable genes that encode human target proteins with ENSG ID for approved drugs or clinical candidates were retained in the following analyses (**Supplementary Table 22**).

We then conducted a series of bioinformatic analyses [i.e., summary-based Mendelian randomization (SMR) and colocalization] to identify prioritized putative druggable genes for BMD treatment. First, we assessed whether the potential genetically regulated expression level of druggable genes were associated with DXA-BMD using SMR ^80^. In SMR analyses, the genetic variants were used as instrumental variables to link the outcome (i.e., DXA-BMD) via the exposure of interest (i.e., the expression level of candidate gene). And the instrumental variables were extracted from the cis-eQTLs in three tissues (i.e., muscle, artery tibial, and whole-blood tissues) from GTEx version 8 projects ^81^ and from eQTLGen consortium (whole blood) ^82^. Linkage clumping was conducted based on default protocols. For each DXA-dervied BMD phenotype, the SMR results of the druggable genes were retained with false discovery rate (FDR)-corrected significance (FDR-corrected *P*_SMR_<0.05 and *P*_HEIDI_>0.05). Among genes with SMR evidence, we further assessed whether the eQTL and DXA-dervied BMD association signals would co-localize at shared loci (i.e., the probability of H4). Specifically, after extracting genetic association estimates of eQTL and DXA-dervied BMD traits with variants within 250kb of the lead SNP, colocalization analyses were performed. The genes with a probability of H4>0.5 were considered to colocalize ^78^. The drug information of genes with SMR evidence was obtained from the GeneCards website (https://www.genecards.org), which collected information from DrugBank, ApexBio, DGIdb, ClinicalTrials.gov, and/or PharmGKB.

## Data Availability

The summary statistics of the present GWASs on 11 DXA-BMD traits and fracture risk were deposited on the WBBC website 83,84 (https://wbbc.westlake.edu.cn/downloads.html). This study does not report the original code.

## Acknowledgments

We thank the High-Performance Computing Center at Westlake University for the facility support and technical assistance. This work was supported by China National GeneBank (CNGB) and KingMed Diagnostics, Co., Ltd.

## Funds

This work was supported by the National Natural Science Foundation of China (#82370887), the “Pioneer” and “Leading Goose” R&D Program of Zhejiang (#2023C03164 and #2024SSYS0032), the Chinese National Key Technology R&D Program, Ministry of Science and Technology (#2021YFC2501702), and the funds from the Westlake Laboratory of Life Sciences and Biomedicine (#202208014).

## Author contributions

H.-F.Z. conceptualized and designed the study. Y.Q. J.X. and P.G. conducted main analysis. L.H., S.X., G.T., H.Y., G.L., and W.X. contributed additional analysis. Y.Q., J.X., and C.X. drafted the manuscript. C.X., S.K., P.Z., M.Y., J.Z., X.L., J.G., S.G., X.Y., J.T., X.C., X.C., and M.M. contributed to the interpretation of the data. P.W., G.L., C.Y., M.Q., Y.F., C.Y., G.C., L.X., D.K., Y.X., L.Y., F.H., C.C., and Z.Z. contributed to reviewing and revising the content of the manuscript. All authors reviewed and approved the final manuscript.

## Data and code availability

The summary statistics of the present GWASs on 11 DXA-BMD traits and fracture risk were deposited on the WBBC website ^83,84^ (https://wbbc.westlake.edu.cn/downloads.html). This study does not report the original code.

