## supplementary figures for "GWAS of ∼30,000 samples with bone mineral density at multiple skeletal sites and its clinical relevance on fracture prediction, genetic correlations and prioritization of drug targets"

### Supplementary Materials

**Supplementary methods**

**Supplementary Figure 1** Overview of this study
**Supplementary Figure 2** Manhattan and quantile-quantile plots for arm BMD.
**Supplementary Figure 3** Manhattan and quantile-quantile plots for femoral neck BMD.
**Supplementary Figure 4** Manhattan and quantile-quantile plots for femur total BMD.
**Supplementary Figure 5** Manhattan and quantile-quantile plots for head BMD.
**Supplementary Figure 6** Manhattan and quantile-quantile plots for leg BMD.
**Supplementary Figure 7** Manhattan and quantile-quantile plots for lumbar spine BMD.
**Supplementary Figure 8** Manhattan and quantile-quantile plots for pelvis BMD.
**Supplementary Figure 9** Manhattan and quantile-quantile plots for rib BMD.
**Supplementary Figure 10** Manhattan and quantile-quantile plots for spine BMD.
**Supplementary Figure 11** Manhattan and quantile-quantile plots for trunk BMD.
**Supplementary Figure 12** Manhattan and quantile-quantile plots for total BMD.
**Supplementary Figure 13** Manhattan and quantile-quantile plots for Fracture.
**Supplementary Figure 14** Regional association plots of flanking 250kb region around the rs746100, based femur total BMD GWAS and *ABCA1* eQTL. The x-axis denotes the physical position of each genetic variant on the chromosome specified, whereas the y-axis indicates the evidence of association, which was shown as $-\log_{10}(P\text{-value})$.
**Supplementary Figure 15** Regional association plots of flanking 250kb region around the rs10840273, based leg BMD GWAS, *SWAP70* eQTL and *SWAP70* pQTL. The x-axis denotes the physical position of each genetic variant on the chromosome specified, whereas the y-axis indicates the evidence of association, which was shown as $-\log_{10}(P\text{-value})$.
**Supplementary Figure 16** Locuszoom of rs12916774 for FNBMD
**Supplementary Figure 17** Locuszoom of rs6013897 for FNBMD
**Supplementary Figure 18** Locuszoom of rs927059 for FNBMD
**Supplementary Figure 19** Cumulative incidence curves for incident fracture across polygenic risk categories in whole population
**Supplementary Figure 20** Cumulative incidence curves for incident fracture across polygenic risk categories in male
**Supplementary Figure 21** Cumulative incidence curves for incident fracture across polygenic risk categories in female

#### Supplementary Methods

#### Gene functional enrichment analysis

For the detected genes, we used KOBAS 3.0 to evaluate the underlying biological mechanisms by using gene ontology (GO) term enrichment analyses [1]. Statistical significance for enrichment analyses was set at an FDR-corrected *P*-value of 0.05.

#### pQTL MR analysis

Based on the SWAP70 pQTL dataset from deCODE, we selected the genome-wide significant independent SNPs across the whole genome as instrumental variables for circulating SWAP70, using PLINK1.90 software (--clump-p1 5e-08, --clump-kb 250, --clump-r2 0.1) [2]. After extraction of genetic association estimates for exposure (i.e., circulating SWAP70) and outcome (i.e., BMD), we performed the inverse-variance weighted method and supplied the weighted median method to assess the association between genetically predicted circulating SWAP70 and BMD.

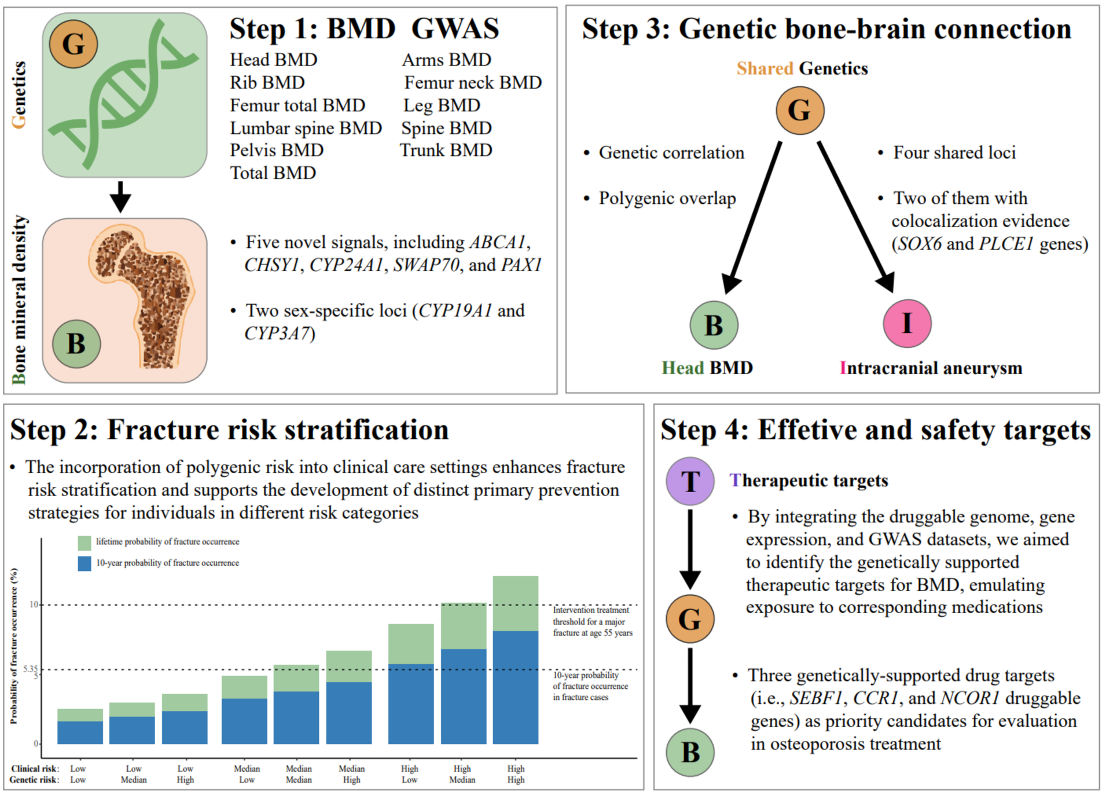


**Supplementary Figure 1** Overview of this study

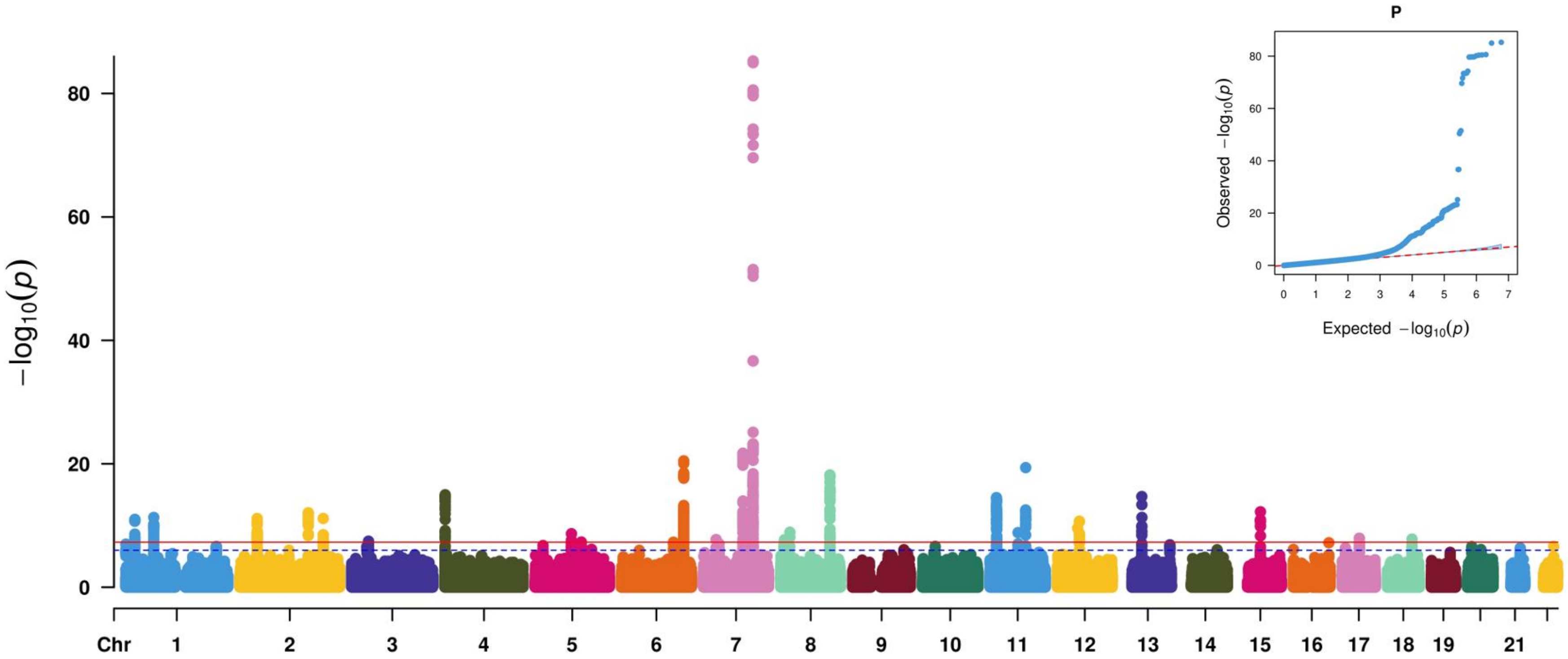


**Supplementary Figure 2** Manhattan and quantile-quantile plots for arm BMD.

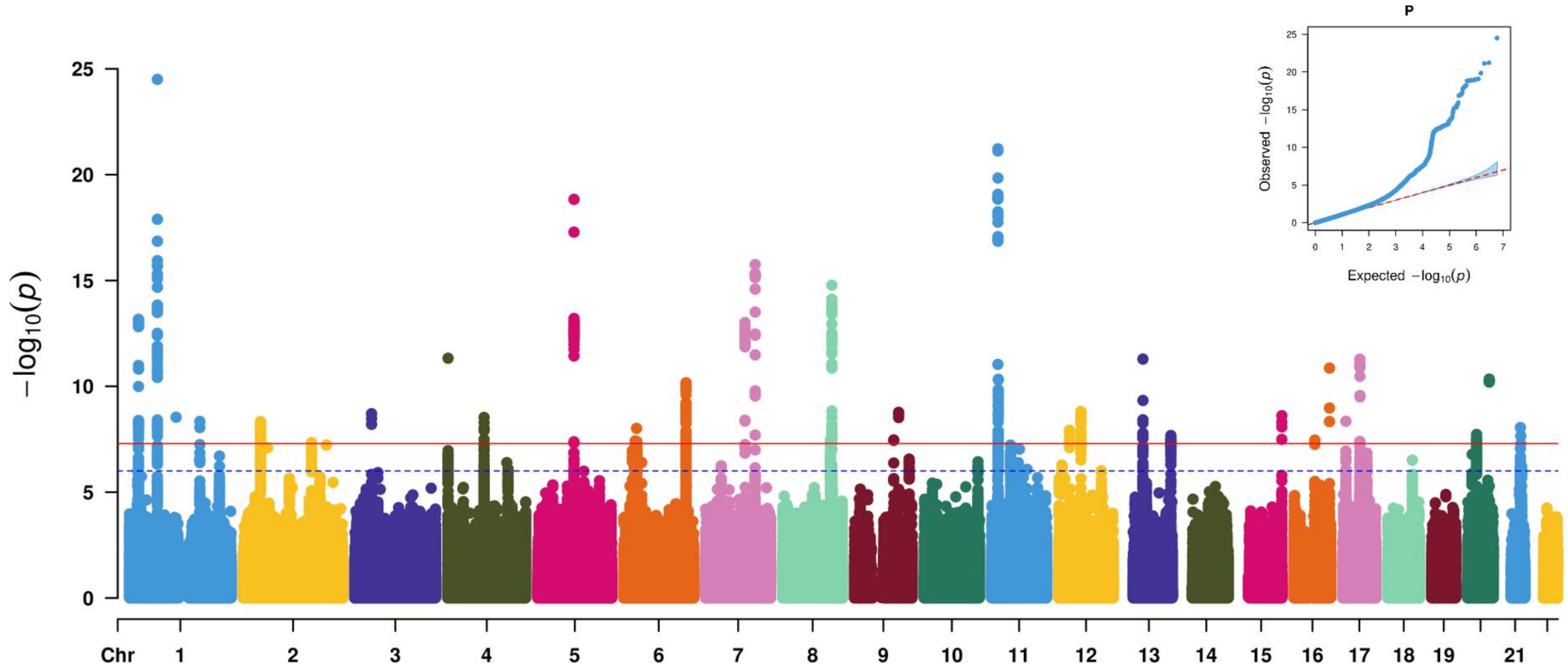


**Supplementary Figure 3** Manhattan and quantile-quantile plots for femoral neck BMD.

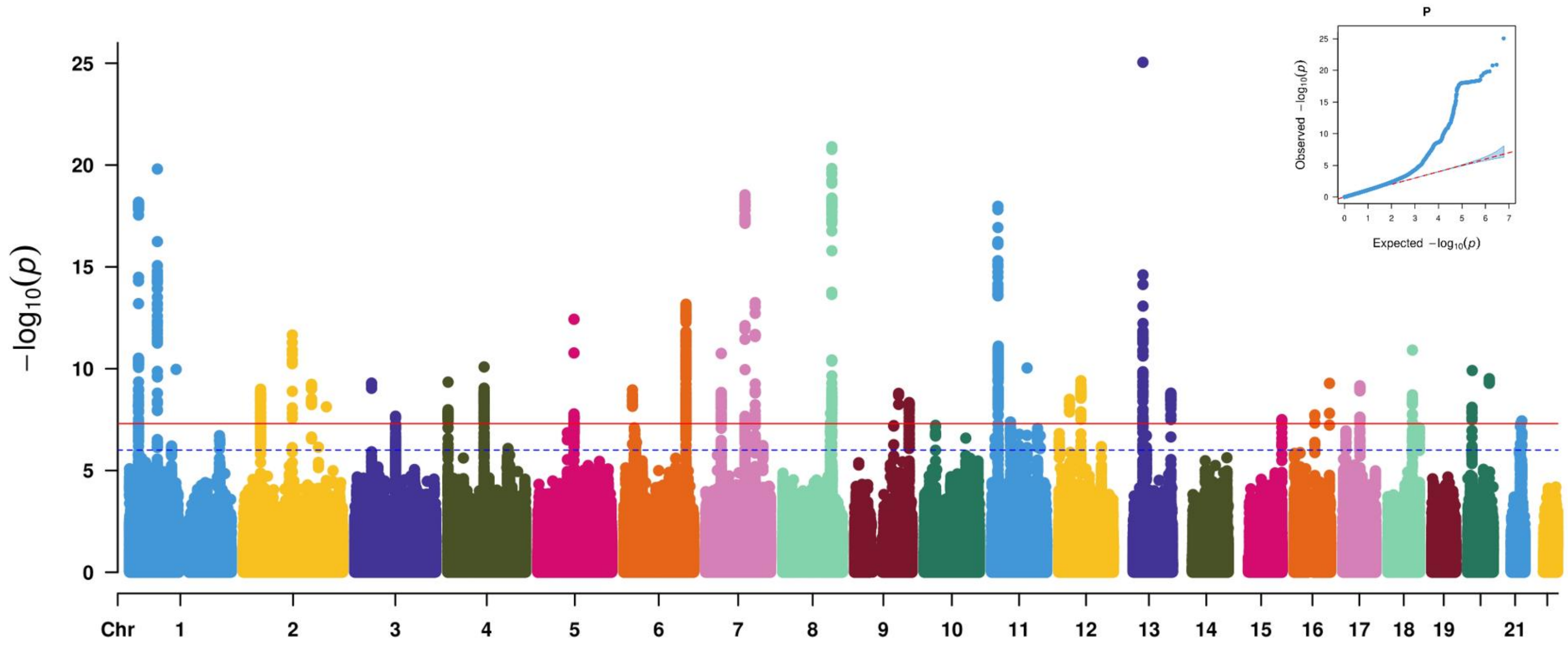


**Supplementary Figure 4** Manhattan and quantile-quantile plots for femur total BMD.

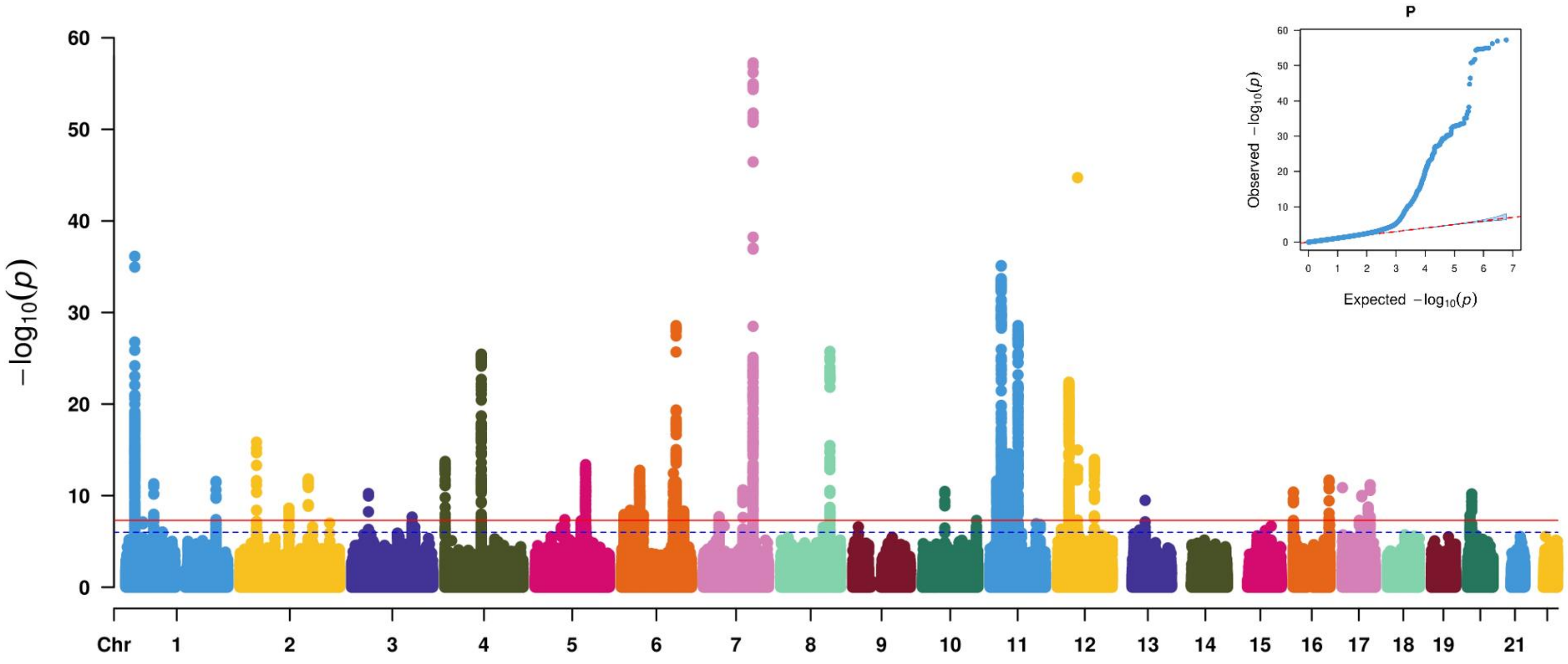


**Supplementary Figure 5** Manhattan and quantile-quantile plots for head BMD.

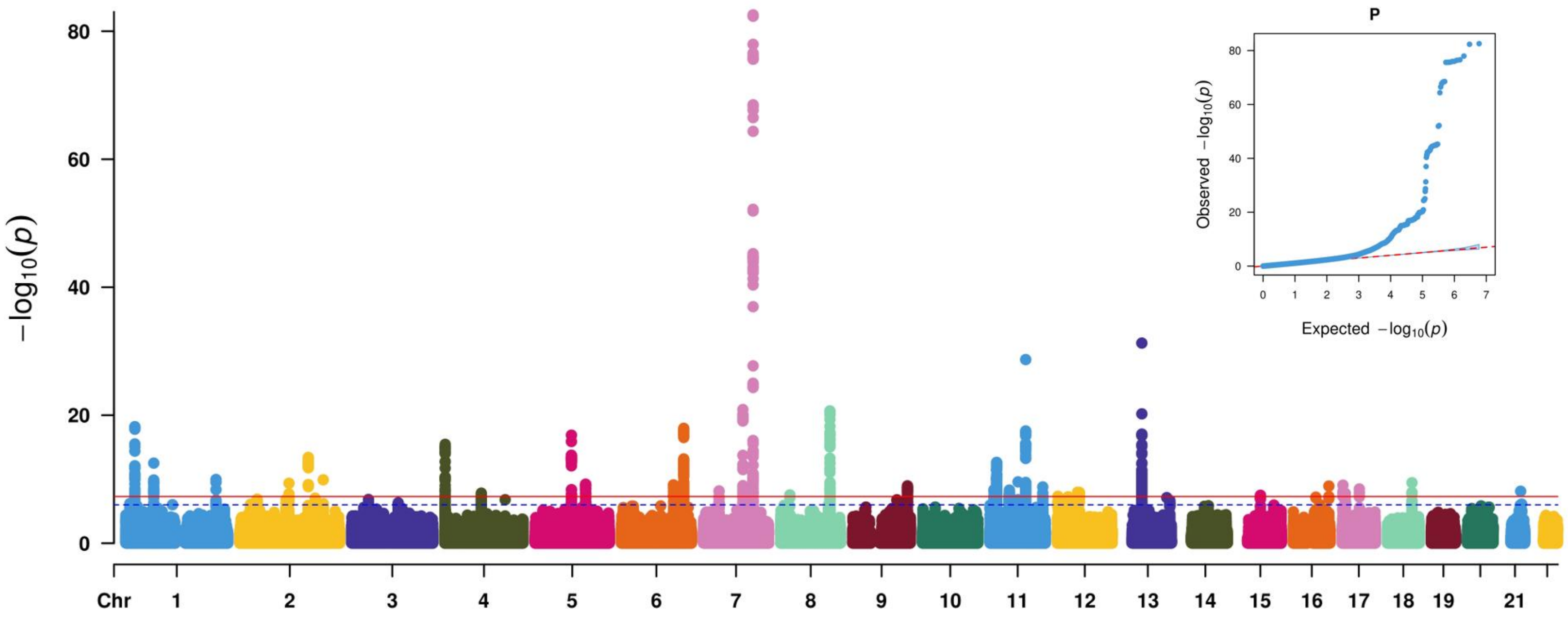


**Supplementary Figure 6** Manhattan and quantile-quantile plots for leg BMD.

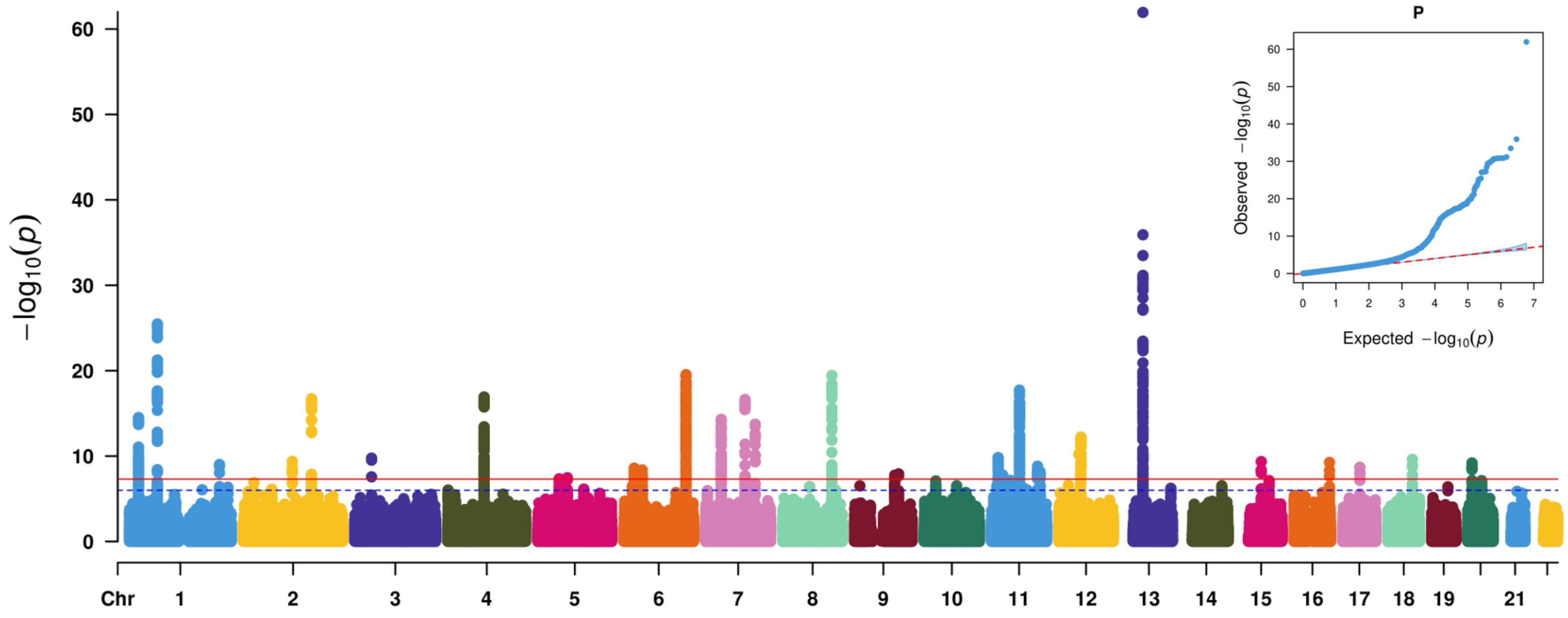


**Supplementary Figure 7** Manhattan and quantile-quantile plots for lumbar spine BMD.

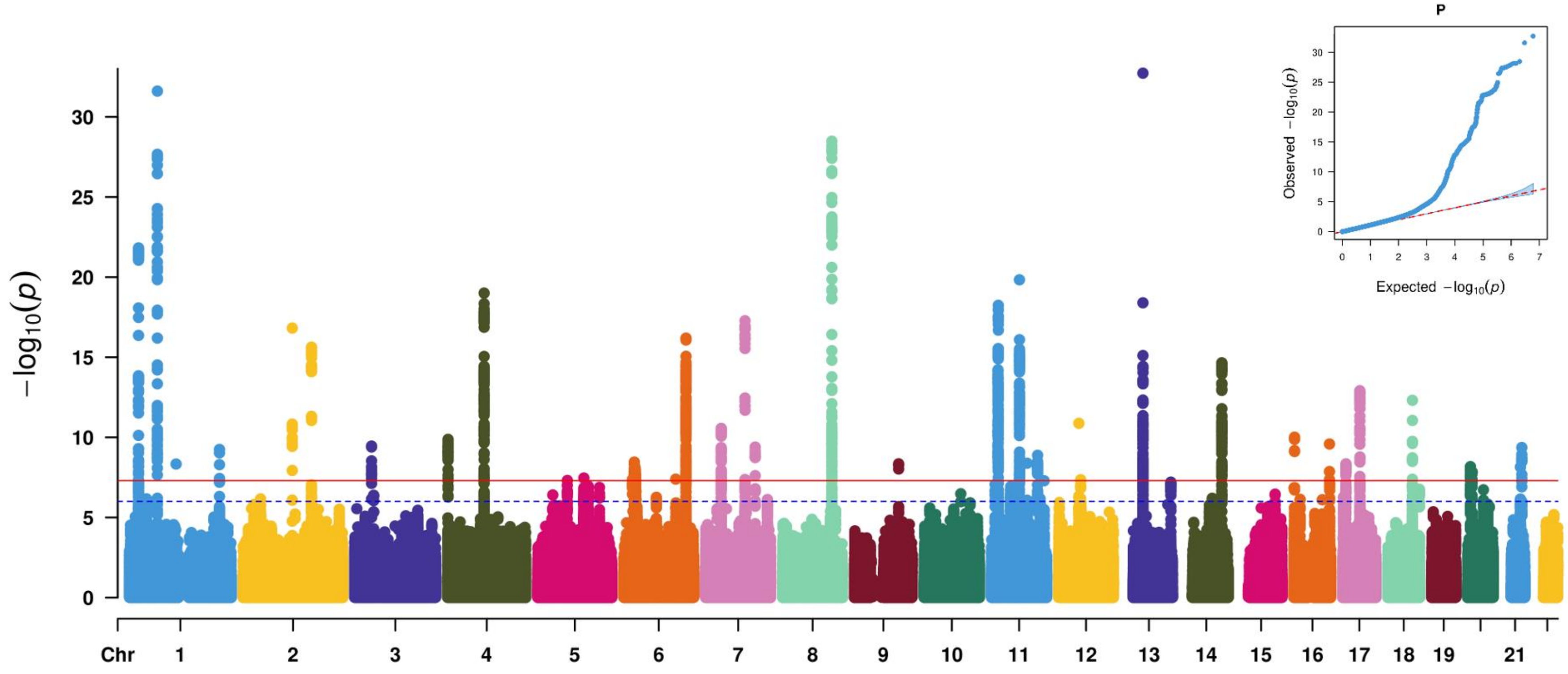


**Supplementary Figure 8** Manhattan and quantile-quantile plots for pelvis BMD.

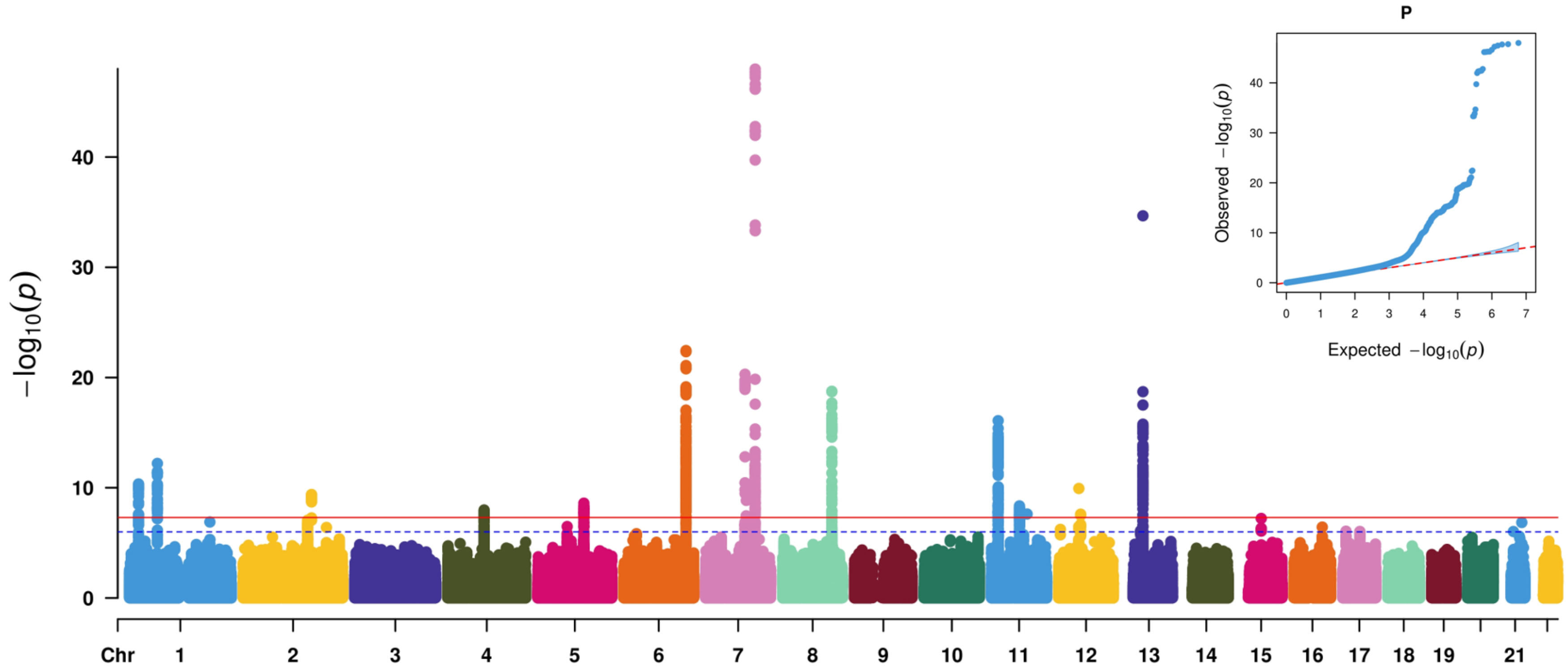


**Supplementary Figure 9** Manhattan and quantile-quantile plots for rib BMD.

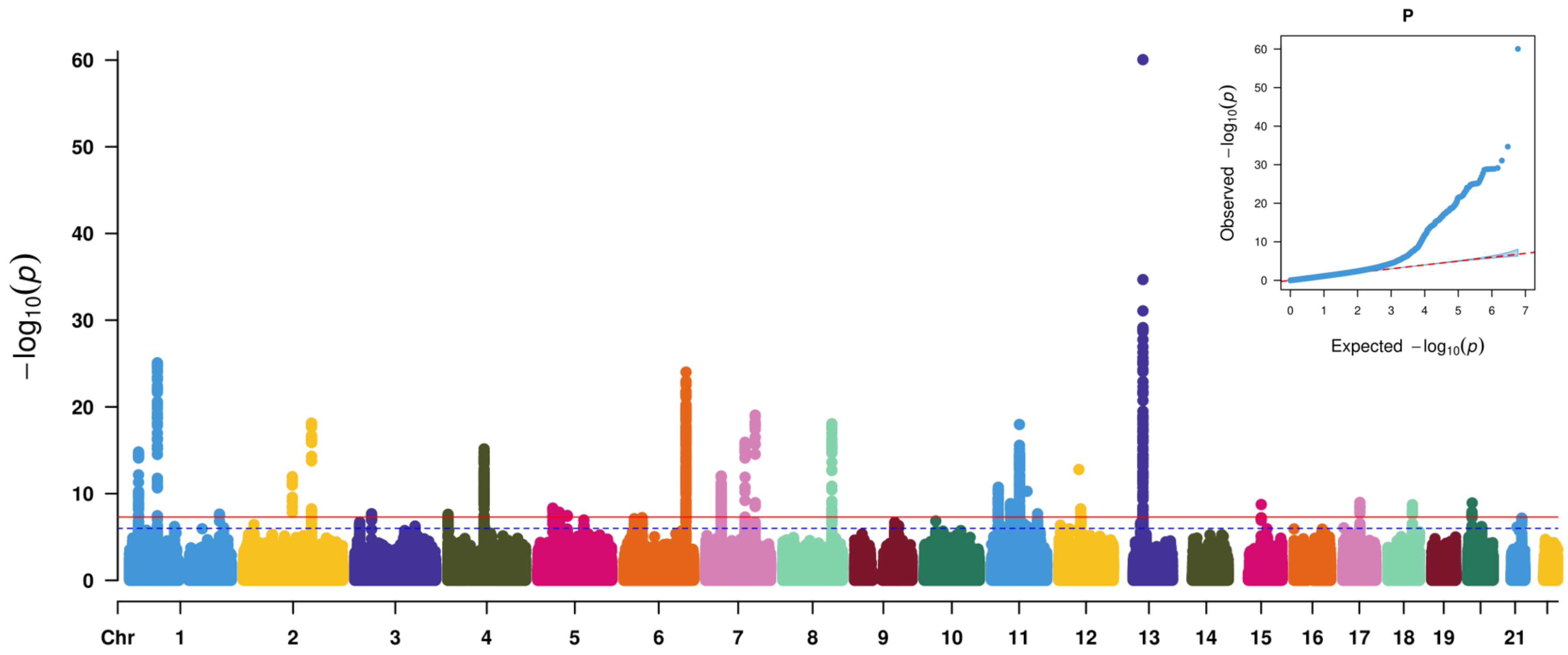


**Supplementary Figure 10** Manhattan and quantile-quantile plots for spine BMD.

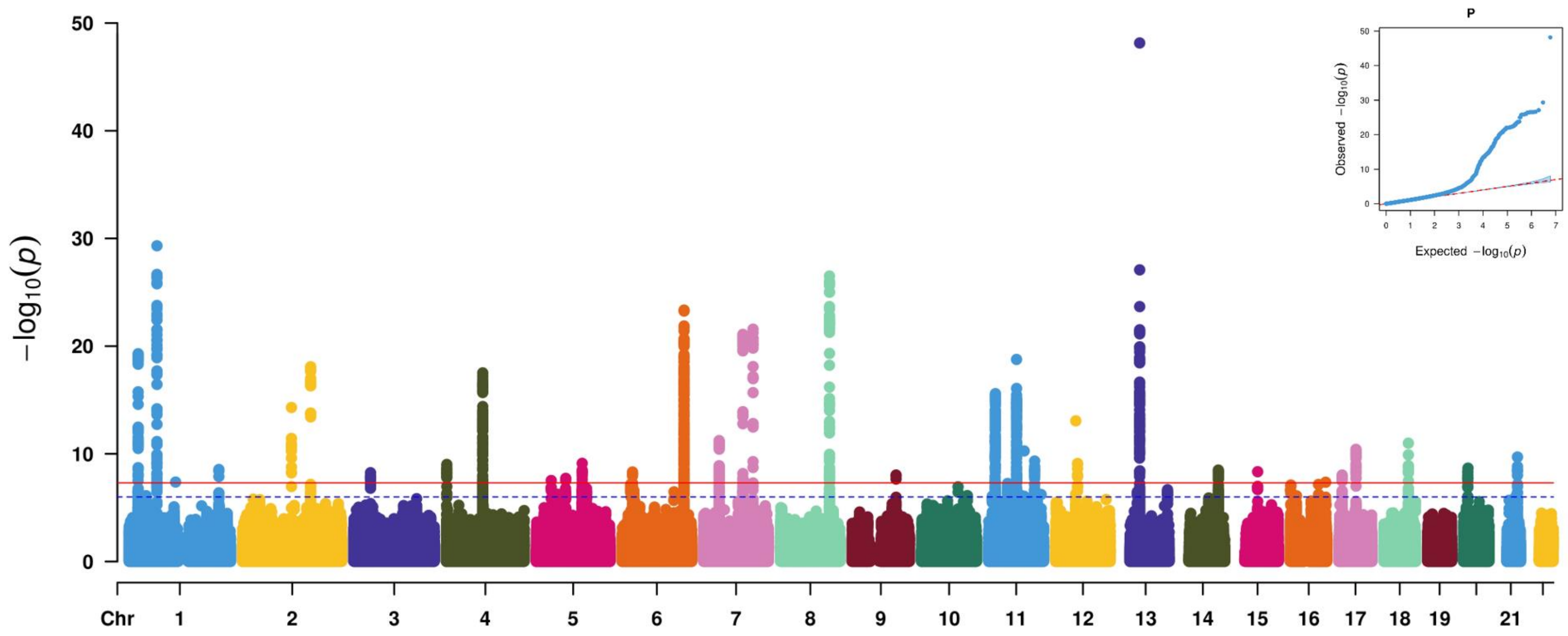


**Supplementary Figure 11** Manhattan and quantile-quantile plots for trunk BMD.

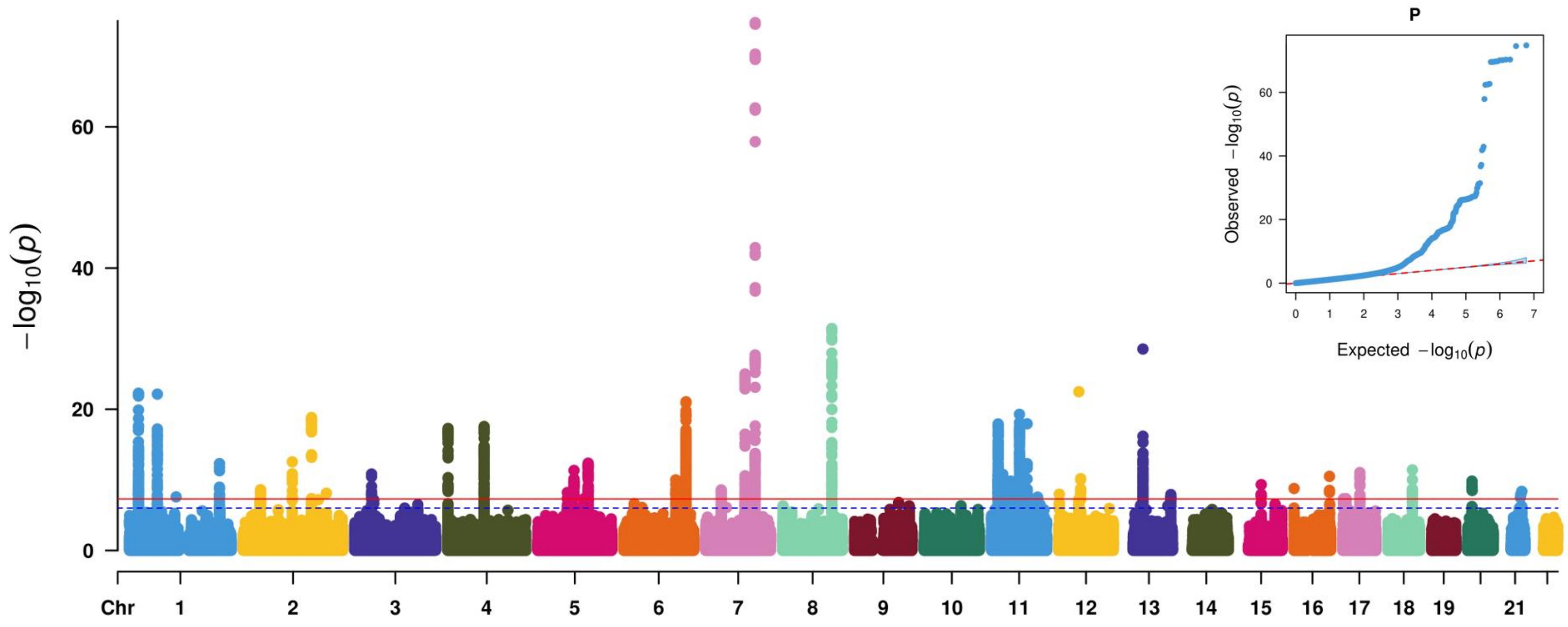


**Supplementary Figure 12** Manhattan and quantile-quantile plots for total BMD.

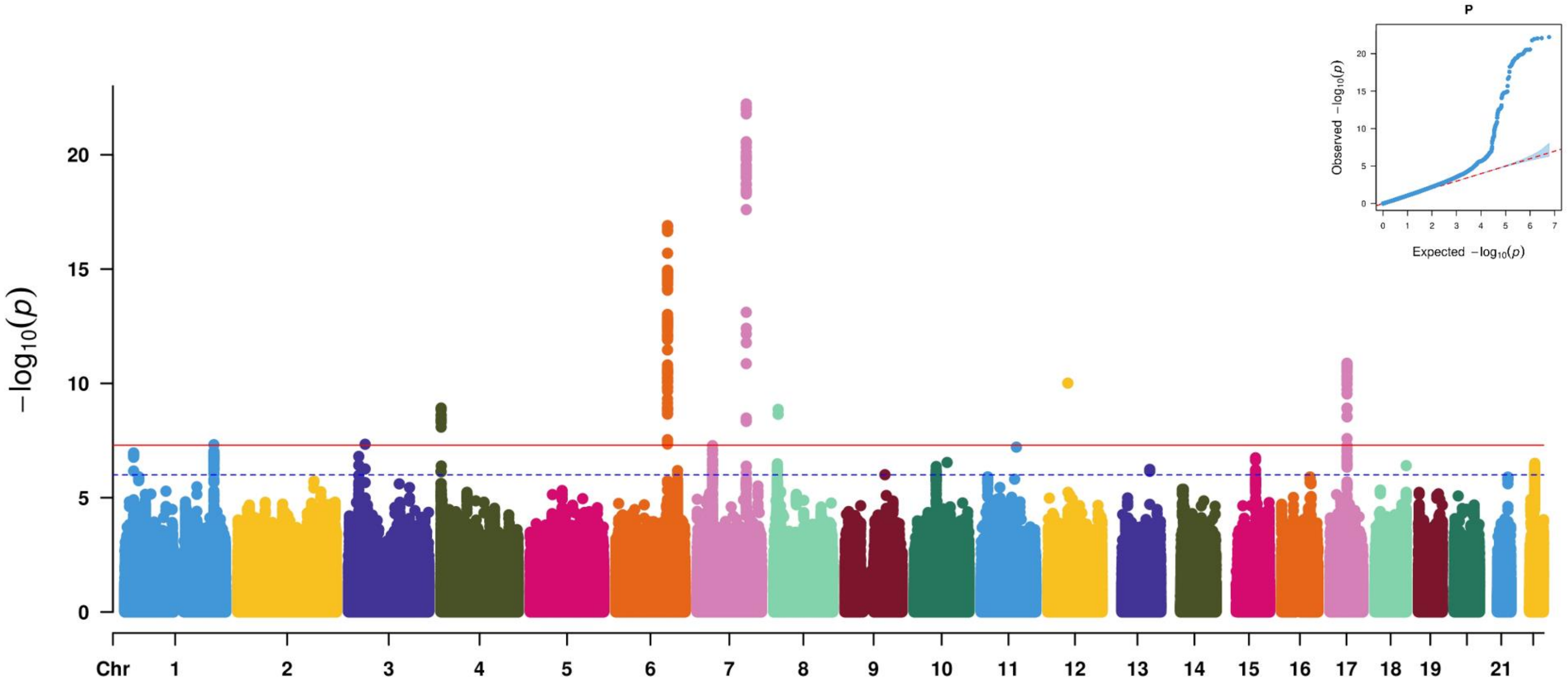


**Supplementary Figure 13** Manhattan and quantile-quantile plots for Fracture.

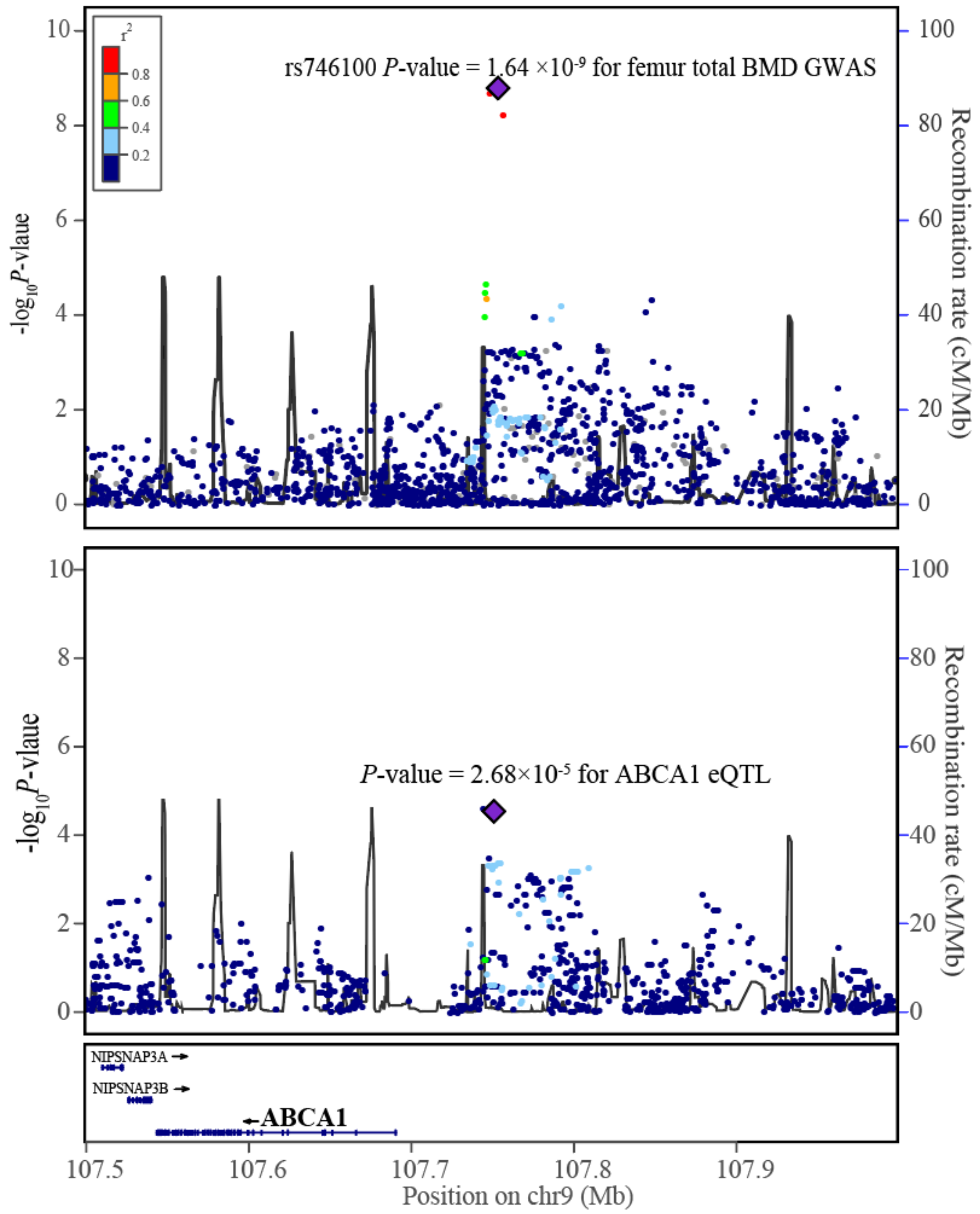


**Supplementary Figure 14** Regional association plots of flanking 250kb region around the rs746100, based femur total BMD GWAS and *ABCA1* eQTL. The x-axis denotes the physical position of each genetic variant on the chromosome specified, whereas the y-axis indicates the evidence of association, which was shown as $-\log_{10}$(*P*-value).

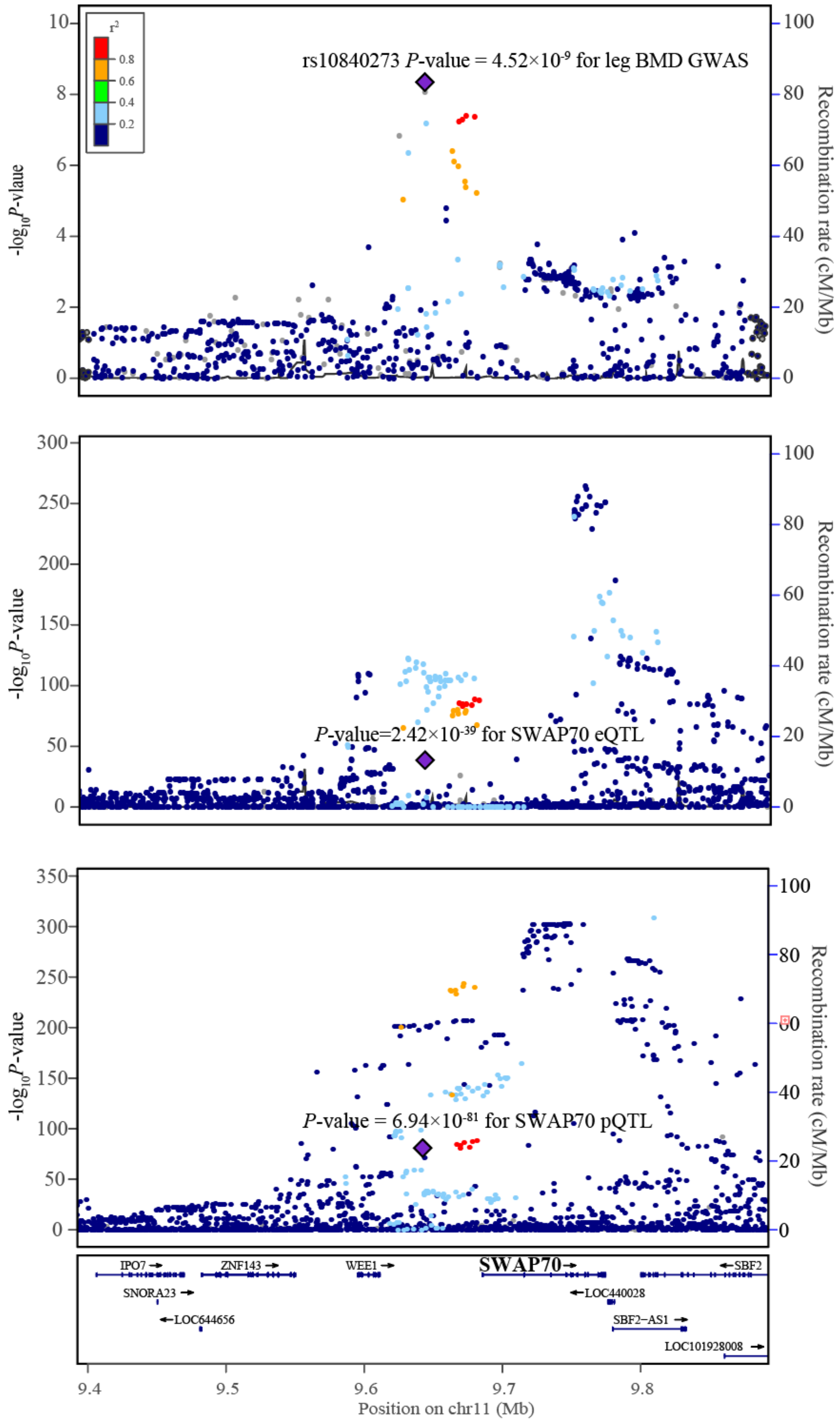


**Supplementary Figure 15** Regional association plots of flanking 250kb region around the rs10840273, based leg BMD GWAS, *SWAP70* eQTL and *SWAP70* pQTL. The x-axis denotes the physical position of each genetic variant on the chromosome specified, whereas the y-axis indicates the evidence of association, which was shown as $-\log_{10}$(*P*-value).

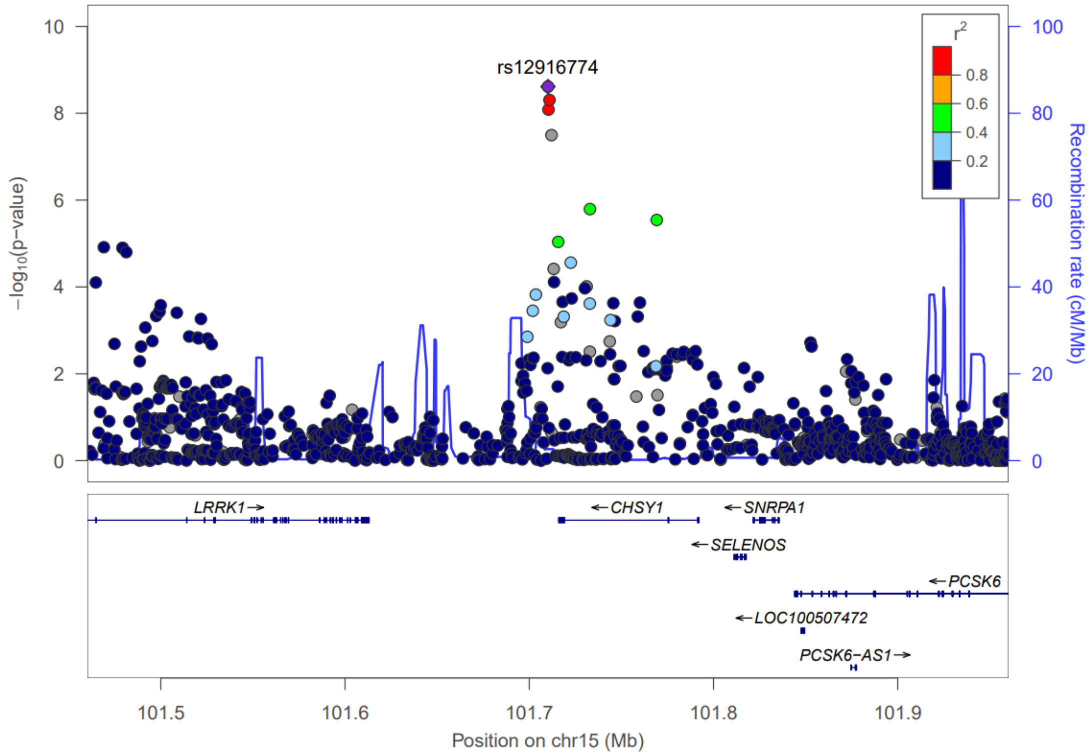


**Supplementary Figure 16** Locuszoom of rs12916774 for FNBMD

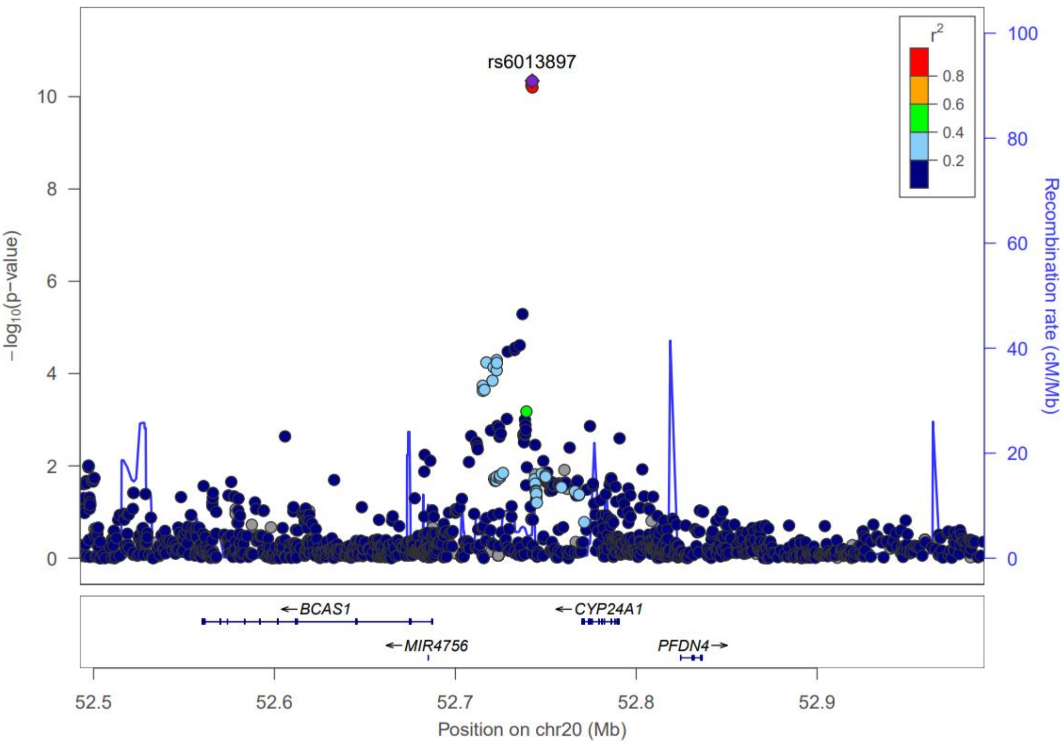


**Supplementary Figure 17** Locuszoom of rs6013897 for FNBMD

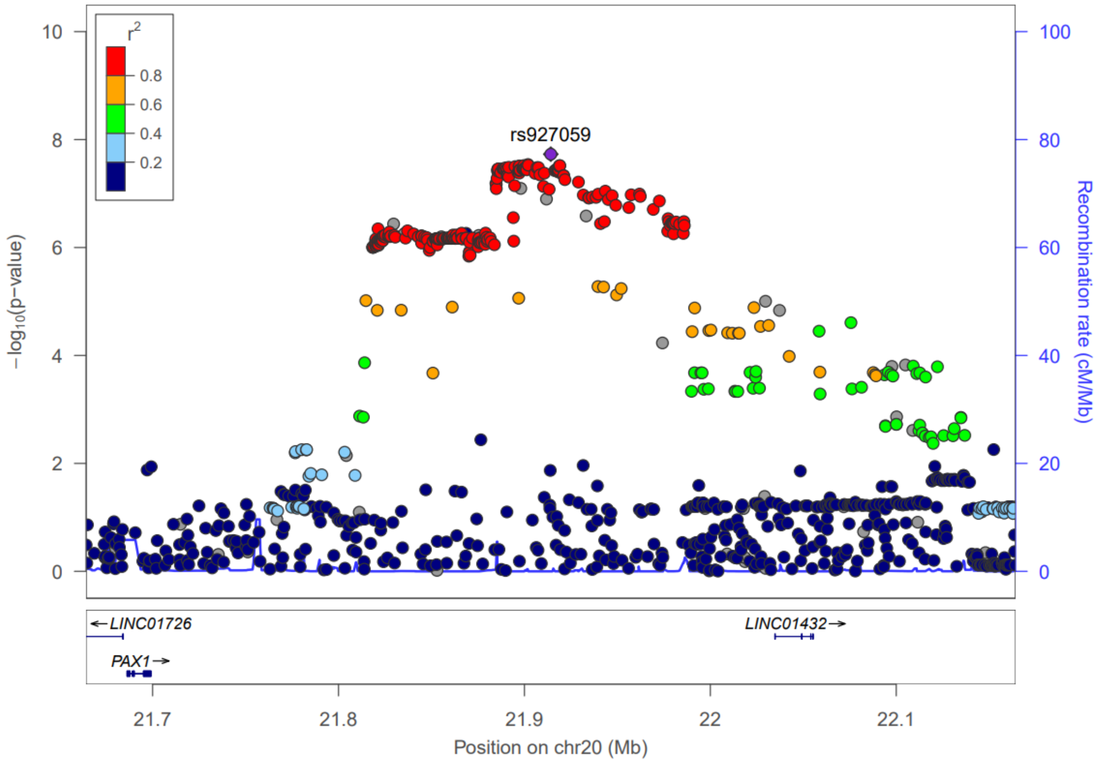


**Supplementary Figure 18** Locuszoom of rs927059 for FNBMD

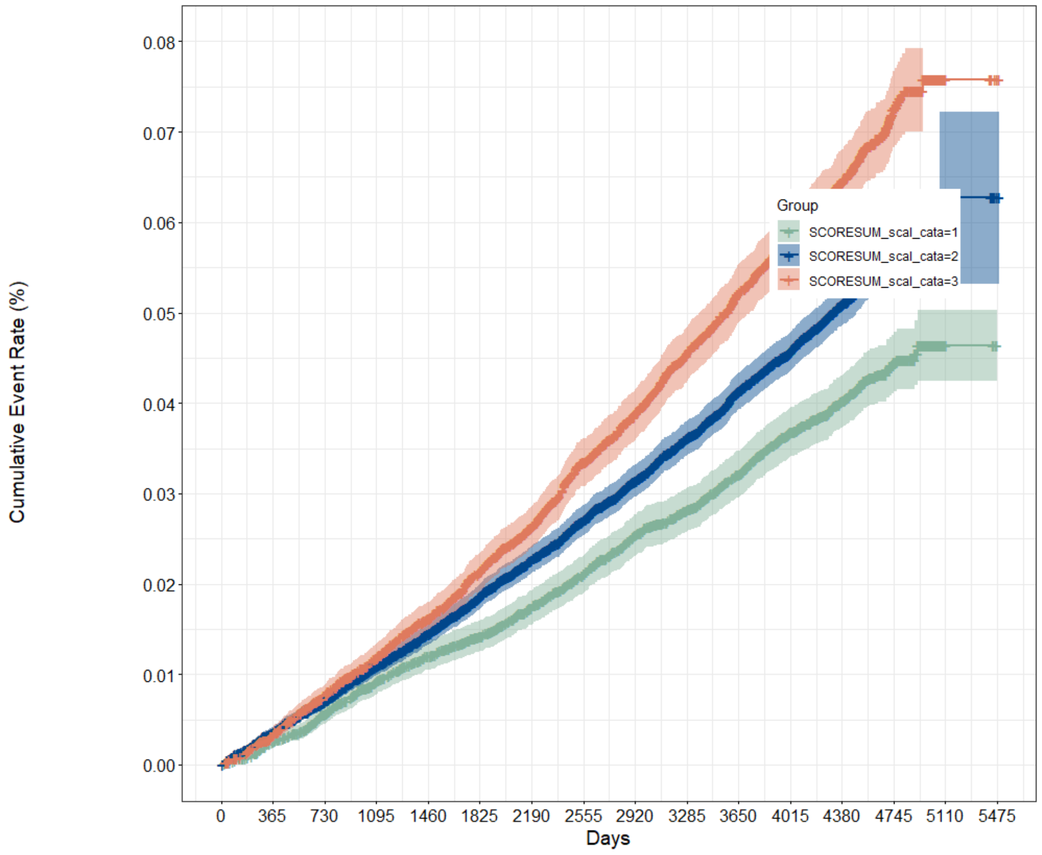


**Supplementary Figure 19** Cumulative incidence curves for incident fracture across polygenic risk categories in whole population

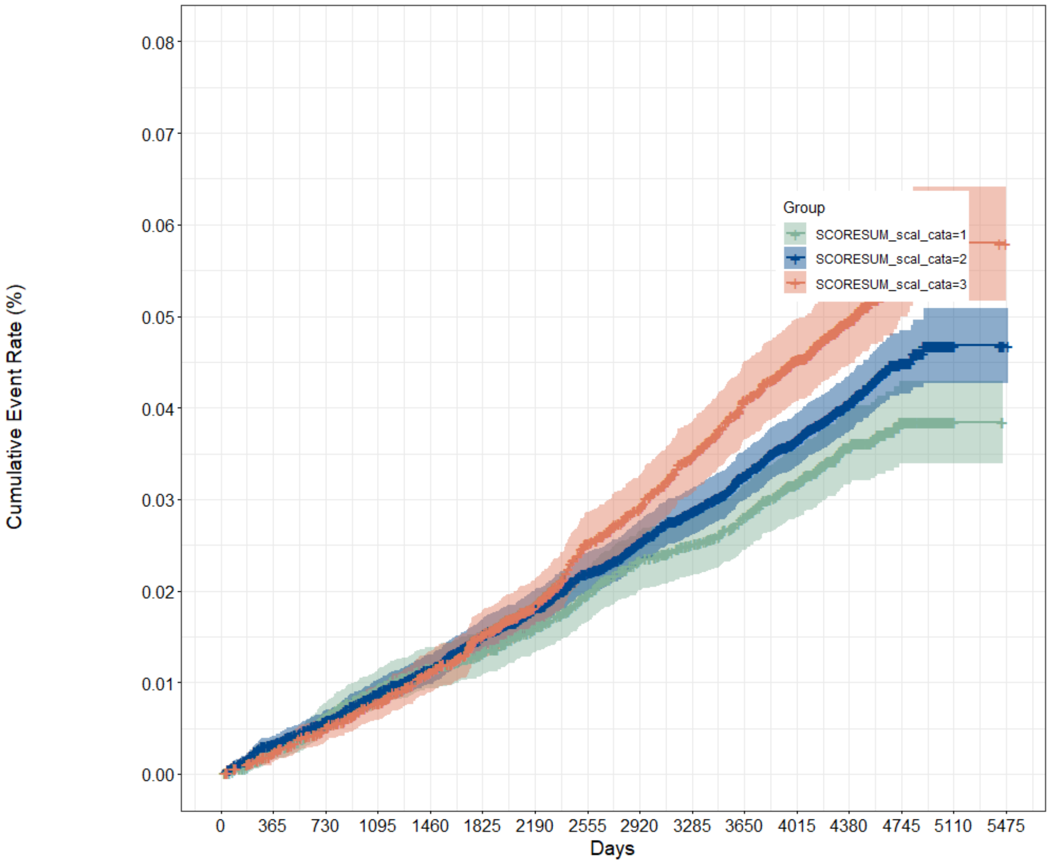


**Supplementary Figure 20** Cumulative incidence curves for incident fracture across polygenic risk categories in male

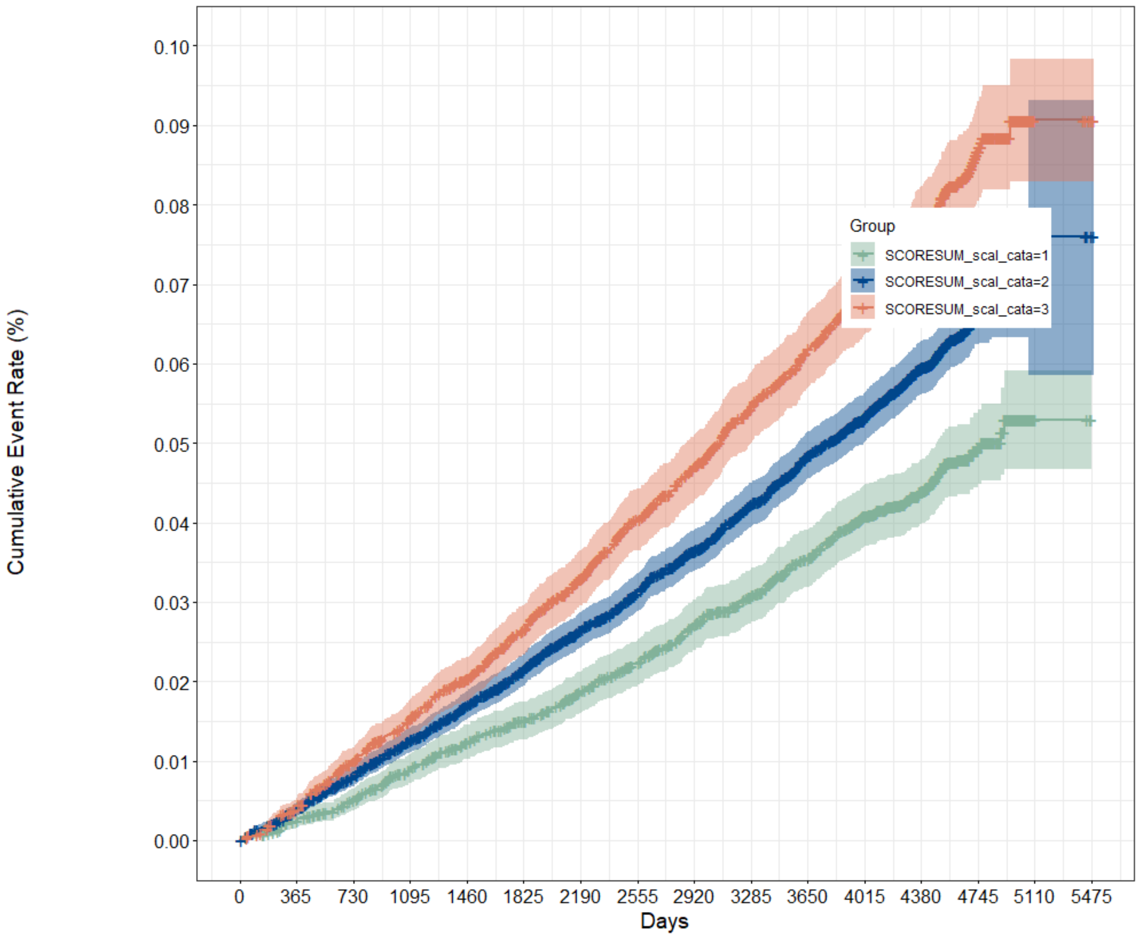


**Supplementary Figure 21** Cumulative incidence curves for incident fracture across polygenic risk categories in female
